# Effects of COVID-19 Mental Health Interventions among Community-based Children, Adolescents, and Adults: A Living Systematic Review of Randomised Controlled Trials

**DOI:** 10.1101/2021.05.04.21256517

**Authors:** Olivia Bonardi, Yutong Wang, Kexin Li, Xiaowen Jiang, Ankur Krishnan, Chen He, Ying Sun, Yin Wu, Jill T. Boruff, Sarah Markham, Danielle B. Rice, Ian Thombs-Vite, Amina Tasleem, Tiffany Dal Santo, Anneke Yao, Marleine Azar, Branka Agic, Christine Fahim, Michael S. Martin, Sanjeev Sockalingam, Gustavo Turecki, Andrea Benedetti, Brett D. Thombs

**Affiliations:** Lady Davis Institute for Medical Research, Jewish General Hospital, Montreal, Quebec, Canada; Department of Psychiatry, McGill University, Montreal, Quebec, Canada; Schulich Library of Physical Sciences, Life Sciences, and Engineering, McGill University, Montreal, Quebec, Canada; Department of Biostatistics and Health Informatics, King’s College London, London, UK; Department of Psychology, McGill University, Montreal, Quebec, Canada; Centre for Addiction and Mental Health, Toronto, Ontario, Canada; Dalla Lana School of Public Health, University of Toronto, Toronto, Ontario, Canada; Li Ka Shing Knowledge Institute, Unity Health Toronto, Toronto, Ontario, Canada; School of Epidemiology and Public Health, University of Ottawa; Ontario, Canada; Correctional Service of Canada, Ottawa, Ontario, Canada; Department of Psychiatry, University of Toronto, Toronto, Ontario, Canada; McGill Group for Suicide Studies, Douglas Mental Health University Institute, McGill University, Montreal, Quebec, Canada; Department of Epidemiology, Biostatistics, and Occupational Health, McGill University, Montreal, Quebec, Canada; Department of Medicine, McGill University, Montreal, Quebec, Canada; Respiratory Epidemiology and Clinical Research Unit, McGill University Health Centre, Montreal, Quebec, Canada; Department of Educational and Counselling Psychology, McGill University, Montreal, Quebec, Canada; Biomedical Ethics Unit, McGill University, Montreal, Quebec, Canada

## Abstract

**Background:** Scalable interventions to address COVID-19 mental health are needed. Our objective was to assess effects of mental health interventions for community-based children, adolescents, and adults.

**Methods:** We searched 9 databases (2 Chinese-language) from December 31, 2019 to March 22, 2021. We included randomised controlled trials with non-hospitalised, non-quarantined participants of interventions to address COVID-19 mental health challenges. We synthesized results descriptively but did not pool quantitatively due to substantial heterogeneity of populations and interventions and concerns about risk of bias.

**Findings:** We identified 9 eligible trials, including 3 well-conducted, well-reported trials that tested interventions designed specifically for COVID-19 mental health challenges, plus 6 trials of standard interventions (e.g., individual or group therapy, expressive writing, mindfulness recordings) minimally adapted for COVID-19, all with risk of bias concerns. Among the 3 COVID-19-specific intervention trials, one (N = 670) found that a self-guided, internet-based cognitive-behavioural intervention targeting dysfunctional COVID-19 worry significantly reduced COVID-19 anxiety (standardized mean difference [SMD] 0.74, 95% CI 0.58 to 0.90) and depression symptoms (SMD 0.38, 95% CI 0.22 to 0.55) in Swedish general population participants. A lay-delivered telephone intervention for homebound older adults in the United States (N = 240) and a peer-moderated education and support intervention for people with a rare autoimmune condition from 12 countries (N = 172) significantly improved anxiety (SMD 0.35, 95% CI 0.09 to 0.60; SMD 0.31, 95% CI 0.03 to 0.58) and depressive symptoms (SMD 0.31, 95% CI 0.05 to 0.56; SMD 0.31, 95% CI 0.07 to 0.55) 6-weeks post-intervention, but these were not significant immediately post-intervention. No trials in children or adolescents were identified.

**Interpretation:** Internet-based programs for the general population and lay-or peer-delivered interventions for vulnerable groups may be effective, scalable options for public mental health in COVID-19. More well-conducted trials, including for children and adolescents, are needed.

**Funding:** Canadian Institutes of Health Research (CMS-171703; MS1-173070); McGill Interdisciplinary Initiative in Infection and Immunity Emergency COVID-19 Research Fund (R2-42).

**Registration:** PROSPERO (CRD42020179703); registered on April 17, 2020.

**RESEARCH IN CONTEXT:** *Evidence before this study:* We searched for systematic reviews of randomised controlled trials of interventions to address mental health challenges in COVID-19. We used searches from our living systematic review, which were not limited by study design and reviewed citations through April 29, 2021 from MEDLINE, PsycINFO, CINAHL, EMBASE, Web of Science, China National Knowledge Infrastructure, Wanfang, medRxiv (preprints), and Open Science Framework Preprints (preprint server aggregator). We identified 4 systematic reviews of interventions for COVID-19 mental health with search dates between April and September 2020. None, however, included evidence from any completed randomised controlled trials on mental health interventions for community-based children, adolescents, or adults during COVID-19.

*Added value of this study:* Our systematic review is the only living systematic review on COVID-19 community-based mental health interventions registered in PROSPERO and, to the best of our knowledge, the first systematic review to synthesize evidence on completed randomised controlled trials of COVID-19 mental health interventions. The sheer volume of evidence being published in COVID-19 poses a barrier to effective synthesis and policy response. We reviewed over 45,000 citations in any language and distilled this to 9 verified eligible community-based trials. Of these, there were 3 well-conducted trials of interventions designed specifically to be scalable to address challenges of public mental health in COVID-19. One trial showed that internet-based cognitive behavioural therapy in the Swedish general population (N = 670) reduced COVID-19 anxiety and symptoms of depression. Trials that tested a lay-delivered telephone support intervention for homebound older adults in the United States (N = 240) and a peer-moderated group intervention for people with a rare autoimmune condition from 12 countries (N = 172) also found that they improved mental health outcomes, although not all outcomes were statistically significant.

*Implications of all the available evidence:* Effective, scalable, and feasibly delivered mental health interventions are needed for the general public and vulnerable groups as lockdown restrictions continue, even intermittently, and because COVID-19 mental health implications will likely persist beyond the pandemic. Although we identified only 3 high-quality trials, they demonstrated approaches that can be feasibly adopted to meet the needs of adults in the general public and vulnerable groups. The successful internet-based cognitive behavioural therapy intervention was made available to the Swedish general public free-of-charge following testing and suggests that online tools tailored for specific concerns in COVID-19 may represent an efficient way of addressing public mental health. Two lay-and peer-delivered interventions, consistent with pre-COVID-19 evidence, suggest that low-intensity, non-professionally delivered, support-oriented approaches can be leveraged among vulnerable groups. The absence of trials of interventions for children and adolescents underlines the need for evidence on scalable strategies for this population, including school-based approaches.

## INTRODUCTION

The SARS-CoV-2 coronavirus disease (COVID-19) pandemic has caused over 3 million deaths worldwide and disrupted social, educational, and economic activities.^1,2^ Internationally, people have faced long periods of lockdown and isolation. There are concerns about effects on mental health,^2-4^ particularly among groups vulnerable to health or social and economic effects of COVID-19, including older individuals; children and adolescents; people with pre-existing medical or mental health conditions; essential services personnel; and individuals marginalized due to poverty, race/ethnicity, or other factors.^5^ Vaccination is underway, but lockdown restrictions will likely continue, at least intermittently, and mental health implications may persist.^2^

COVID-19 mental health challenges may include loneliness, boredom, grief and loss, depression, stress, worry, fear, burnout, and anxiety.^2-8^ Scalable mental health interventions, which are interventions that can be feasibly delivered to large numbers of people affected by adversity, are needed.^9^ These could include non-specialist-delivered or self-help versions of evidence-based interventions; guided, group-based interventions; or peer-support interventions, for example.^9,10^

We identified four systematic reviews^11-14^ that have attempted to synthesize evidence on mental health interventions for non-hospitalised children, adolescents, or adults in COVID-19, but all were done early in the pandemic, and none included any randomised controlled trials (RCTs). End dates of searches were between April and September 2020 with none ongoing.

Living systematic reviews15 are systematic reviews that are continually updated to provide up-to-date evidence. They are logistically challenging but highly valuable when (1) important decisions to be made merit the resources involved; (2) low-certainty in existing evidence poses a barrier to decision-making; and (3) new emerging evidence may inform decisions.^15^ Timely evidence is needed to support mental health responses to COVID-19.

We are conducting living systematic reviews^3,4^ of changes in mental health symptoms during COVID-19 and effects of mental health interventions, both for people with COVID-19 infection or exposure and for community-based public mental health. The objective of the present report is to synthesize evidence from RCTs on mental health interventions for community-based children, adolescents, and adults.^3,4^

## METHODS

Our systematic review was registered in the PROSPERO prospective register of systematic reviews (CRD 42020179703), and a protocol was uploaded to the Open Science Framework (https://osf.io/96csg/) prior to initiation. Results are reported in accordance with the PRISMA statement.^16^ Results are also posted online (https://www.depressd.ca/research-question-3-intervention). The present report includes RCTs of community-based mental health interventions, which is a subset of trials included in our main systematic review of interventions.

### Eligible Studies

Our main systematic review of interventions included randomised or non-randomised trials of mental health interventions conducted in any population during COVID-19. All participants had to be enrolled after December 31, 2019, when China first reported on COVID-19 to the World Health Organization.^17^ Eligible interventions included any intervention described as designed to address COVID-19 mental health challenges or primarily addressing mental health symptoms from COVID-19. Trials that were not mental health interventions and primarily targeted non-mental health outcomes (e.g., exercise with primary outcome physical activity) were excluded, even if mental health outcomes were reported. Eligible comparators included: (1) inactive control conditions (e.g., no treatment, waitlist) and (2) other eligible interventions. Eligible outcomes were defined broadly and included general mental health, mental health quality of life, anxiety symptoms, depression symptoms, stress, loneliness, anger, grief, burnout, and other emotional states. To be eligible, trials had to report outcomes collected at least one week after intervention initiation and include at least 10 total participants. There were no restrictions on language or publication format.

The present report focuses on identifying scalable interventions to address public mental health. Thus, it does not include (1) trials done exclusively with hospitalised patients or persons quarantined due to COVID-19 infections or exposure, because they face different challenges than people in the community; (2) trials of brief single-session interventions (e.g., 30 minutes) with no subsequent follow-up, which would not likely inform community mental health programming; or (3) non-randomised studies, because those we identified included few subjects and were not adequately reported. Those non-included trials are shown in the appendices.

Additionally, the high volume of poor-quality research being published on COVID-19 is a barrier to synthesis,^18^ and we encountered many trials that were of extremely poor quality, of unclear origin and sponsorship, and reported effect sizes that, in some cases, exceeded plausibility. Thus, we contacted authors of all included studies by email up to 2 times and requested that they verify the authenticity of published methods and results and confirm the accuracy of our extracted data. Authors of studies published in Chinese-language journals were contacted with text in both English and Chinese. We did not include unverified trials in our main report but instead show results in the appendices.

### Identification and Selection of Eligible Studies

The same search strategies were used for all research questions in our systematic reviews. We searched MEDLINE (Ovid), PsycINFO (Ovid), CINAHL (EBSCO), EMBASE (Ovid), Web of Science Core Collection: Citation Indexes, China National Knowledge Infrastructure, Wanfang, medRxiv (preprints), and Open Science Framework Preprints (preprint server aggregator) using a strategy designed and built by an experienced health sciences librarian. The China National Knowledge Infrastructure and Wanfang databases were searched using Chinese search terms based on the English-language search strategy. The rapid project launch did not allow for formal peer review, but COVID-19 terms were developed in collaboration with other librarians working on the topic. See Appendix 1. Our initial search was conducted from December 31, 2019 to April 13, 2020, then automated searches were set for daily updates. On December 28, 2020, we converted to weekly updates to improve processing efficiency.

Search results were downloaded into the systematic review software DistillerSR (Evidence Partners, Ottawa, Canada), where duplicate references were identified and removed. Two independent reviewers evaluated titles and abstracts in random order. If either reviewer deemed a study potentially eligible, a full-text review was completed, also by two independent reviewers. Discrepancies at the full-text level were resolved through consensus, with a third investigator consulted as necessary. To ensure accurate identification of eligible studies, a coding guide with inclusion and exclusion criteria was developed and pre-tested, and all team members were trained over several sessions. See Appendix 2.

### Data Extraction and Synthesis

For each included study, one reviewer extracted data using a pre-specified standardized form, and a second reviewer validated extracted data. Reviewers extracted (1) publication characteristics (e.g., first author, journal); (2) population characteristics (e.g., country, eligibility criteria, recruitment method, number of participants, age, sex or gender); (3) COVID-19 characteristics (e.g., time during pandemic); (4) intervention components; (5) mental health outcomes; (6) risk of bias; and (7) adequacy of intervention reporting. If sufficient information to calculate effect sizes for one or more outcomes was not provided, we contacted authors to obtain missing information.

We used the 2011 version of the Cochrane Collaboration risk of bias tool.^19^ The tool has 7 domains, including random sequence generation, allocation concealment, blinding of participants and personnel, blinding of outcome assessment, incomplete outcome data, selective outcome reporting, and other sources of bias. Studies were rated low, unclear, or high risk on each domain.

We used the Template for Intervention Description and Replication (TIDieR) checklist to evaluate the degree that interventions were reported adequately for replication in research or practice.^20^ The checklist is comprised of 12 items that assess reporting of intervention name; rationale or theory underlying the intervention; physical or informational material used; procedures and processes; provider and background; delivery mode (e.g., group, face-to-face); location where delivered and necessary infrastructure; number of sessions, schedule, and duration; if tailoring was done and how; any modifications made; if adherence or fidelity was assessed and how; and, if assessed, the extent to which the intervention was delivered as planned.

For included trials, if not provided, we calculated between-groups standardized mean difference (SMDs) using Hedges’ g with 95% confidence intervals (CI).^21^ We did not pool results across trials because of substantial heterogeneity of populations, interventions, and outcomes and concerns about risk of bias. Instead, we reported results descriptively.

### Protocol Amendments

Our systematic review was quickly designed and initiated in April 2020. Several amendments or clarifications were made subsequently. First, we changed from daily to weekly search updates on December 28, 2020 for more efficient reference processing. Second, on January 27, 2021 we made a minor change to search strategies to incorporate a new physical distancing subject heading created for COVID-19. Third, we made several amendments to Chinese-language search strategies to facilitate processing (see Appendix 1). Fourth, we added the TIDieR20 checklist to assess intervention reporting quality. Fifth, we clarified that we only included trials which initiated participant enrolment after December 31, 2019. Sixth, we clarified criteria for assessing whether an intervention addressed mental health related to COVID-19; see Appendix 2. Seventh, we decided to separately report trials addressing community-based mental health and trials for people infected with COVID-19 or quarantined due to exposure, due to major differences in challenges faced by these groups and intervention approaches. Eighth, because we have encountered many trial reports of poor quality with seemingly implausible results, this report only includes trials whose authors verified accuracy of their report and our extracted data; results of other trials are included in appendices.

## RESULTS

### Search Results and Selection of Eligible Studies

As of March 22, 2021, our searches identified 45,777 unique citations. Of these, 45,536 were excluded after title and abstract review and 146 after full-text review, leaving 95 trials, of which 59 evaluated interventions for people hospitalised or quarantined due to COVID-19, 10 assessed single-session interventions without subsequent follow-up, 4 were non-randomised trials, and 13 were not verified by authors (6 without author contact information in publication or online; 7 no response), leaving 9 eligible, verified RCTs for inclusion^22-30^ (Figure 1).

**Figure 1.**
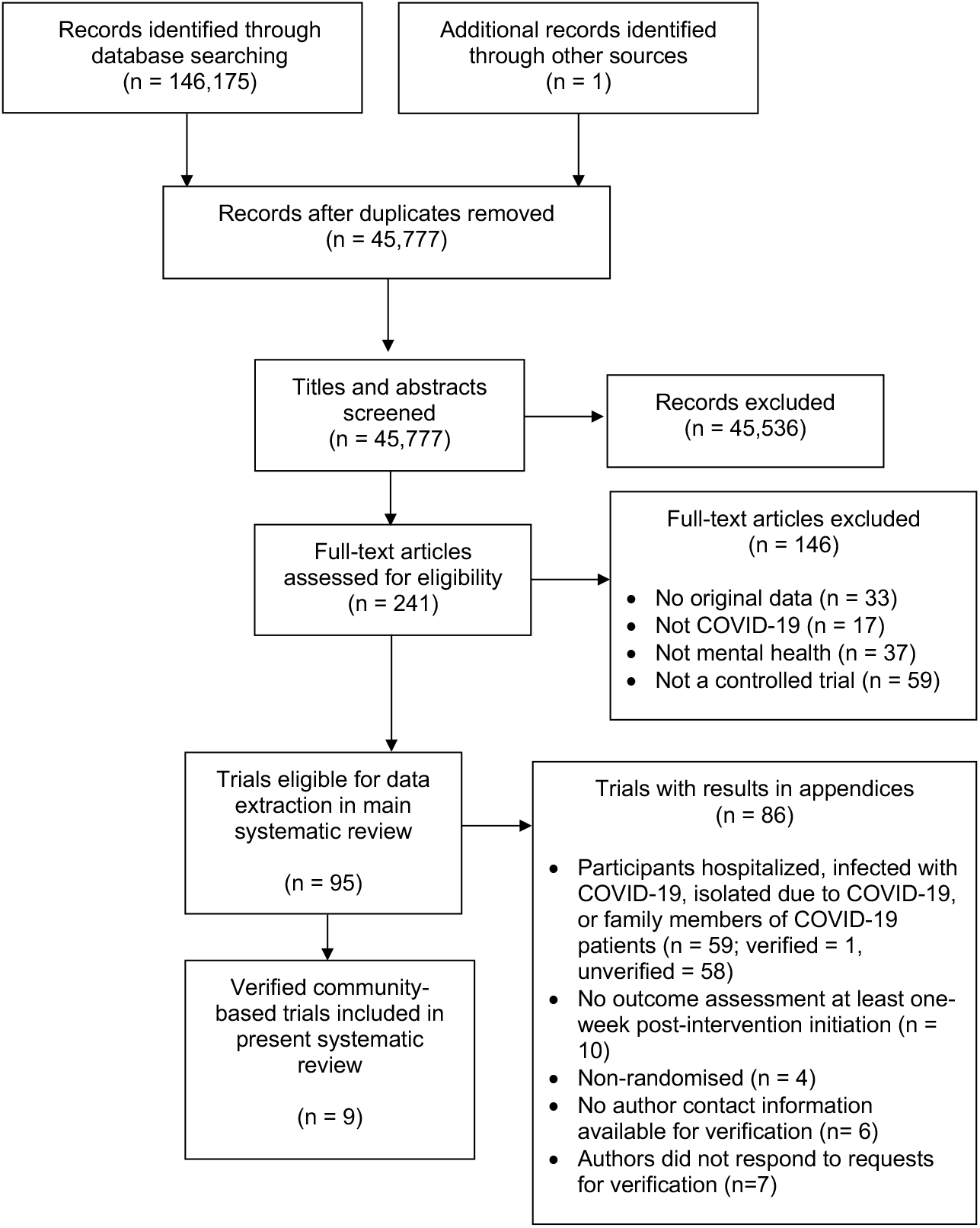
PRISMA 2020 Flow Diagram: Searches through March 22, 2021.

### Characteristics of Included Trials

Table 1 shows the characteristics of included RCTs. See Appendix 3 for characteristics (plus outcomes, risk of bias, intervention reporting) of otherwise eligible but unverified trials and Appendix 4 for trials with hospitalised or quarantined individuals, trials of brief interventions without follow-up, and non-randomised trials. Of the 9 included trials, 3 trials^22-24^ tested interventions designed specifically to address mental health challenges in COVID-19, and 625^-30^ tested standard interventions that were only minimally adapted.

**Table 1.**
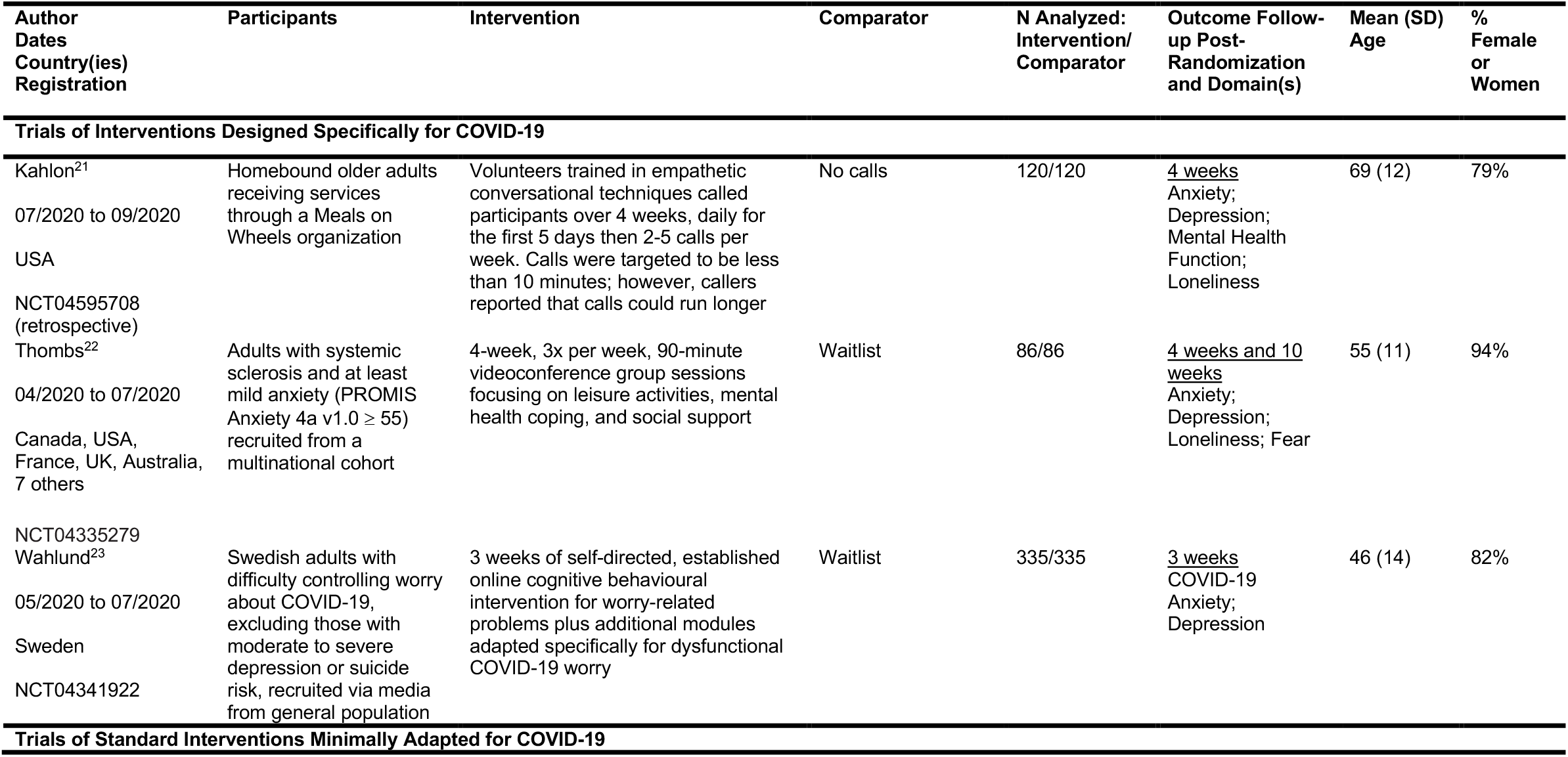
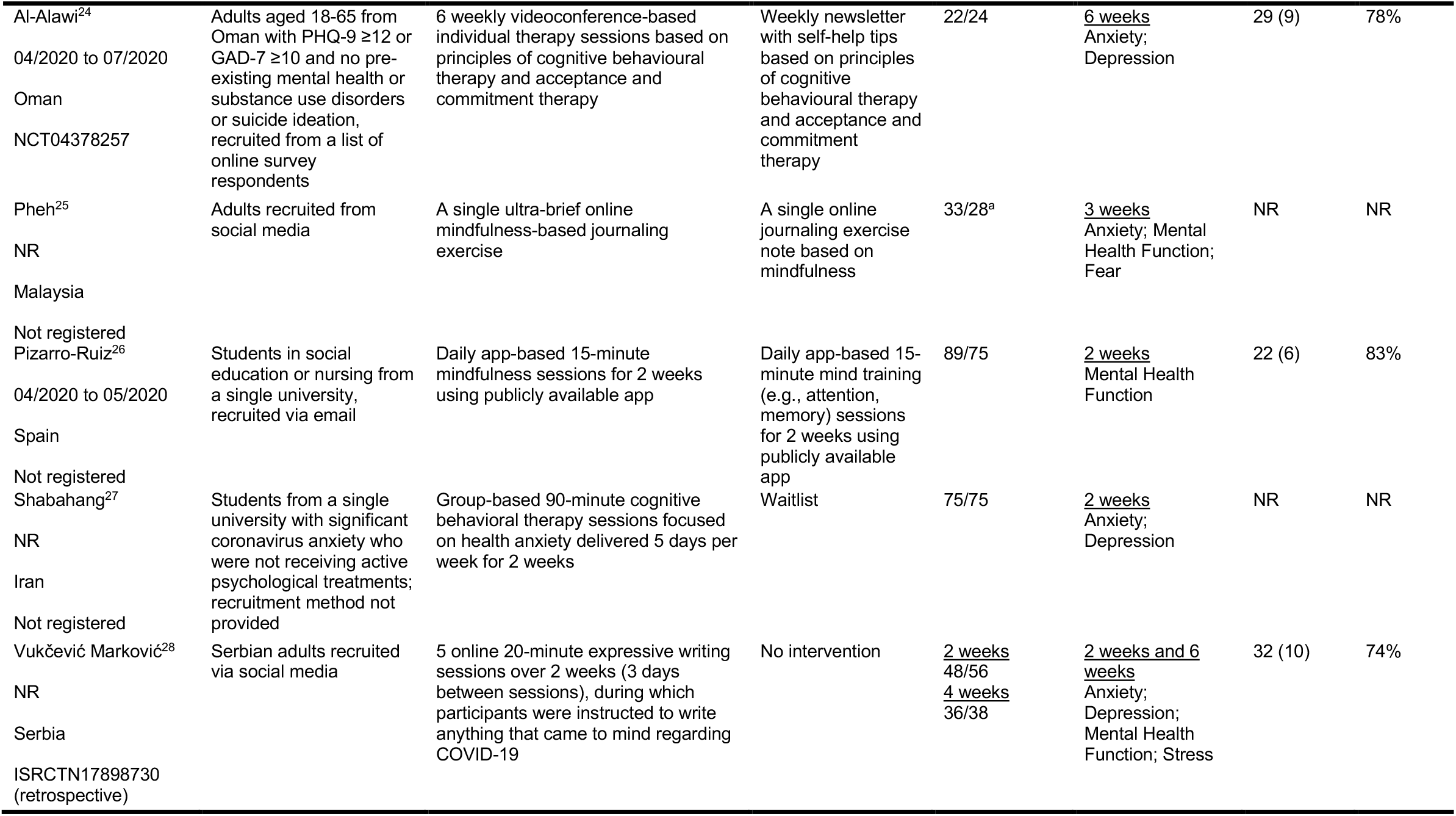
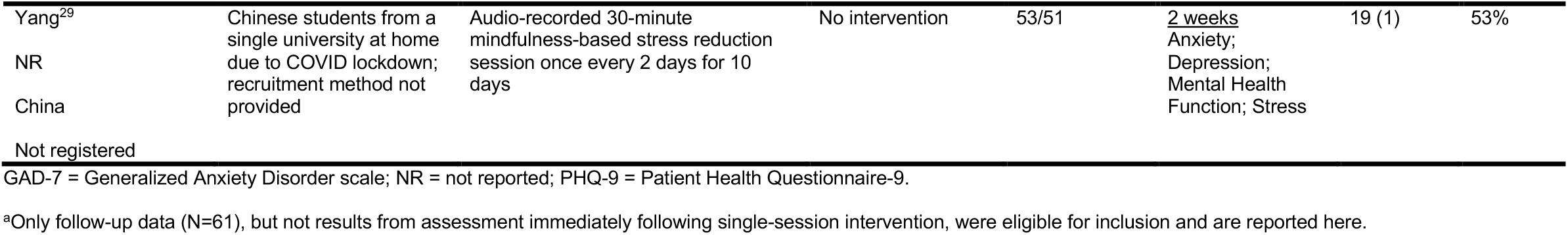
Characteristics of Included Trials.

#### Trials of Interventions Designed Specifically for COVID-19

Among the 3 trials^22-24^ of interventions designed to address mental health challenges in COVID-19, two^22,23^ used lay-delivered or peer-support interventions with groups of vulnerable individuals. The third24 used an online cognitive behavioural therapy intervention to address COVID-19 worry in the general population.

A trial from the United States (N = 240)^22^ tested the effects of 4 weeks of layperson-delivered telephone calls to racially and ethnically diverse homebound older adults receiving home meal services through a Meals on Wheels organization (mean [SD] age = 69 [12], 79% women, 100% chronic medical condition) on anxiety, depressive symptoms, general mental health function, and loneliness. The investigators trained university students in empathetic conversational skills (e.g., prioritizing listening, eliciting conversation on topics of interest to participants), and each caller supported 6 to 9 participants. Calls, which were targeted to be < 10 minutes, were done on 5 days in the first week and 2 to 5 days in the following three weeks.

A second trial^23^ (N = 172) randomized people with the rare autoimmune disease systemic sclerosis, or scleroderma, from 12 countries to receive 4-weeks (3 times per week) of a multi-faceted group videoconference-based intervention or waitlist control. It tested effects of the intervention, which combined activity engagement, education and practice in mental health coping strategies, and peer support on outcomes that included anxiety, depressive symptoms, fear, and loneliness. Groups included 6 to 10 participants and were moderated by peers previously trained as support group leaders.

The third trial (N = 670)^24^ tested effects on COVID-19-related anxiety and depressive symptoms after receiving 3 weeks of access to a self-guided online cognitive behavioural intervention. Adults in the Swedish general population were recruited through advertising on national television, newspapers, and social media and randomised to the intervention or waitlist control. The intervention was based on established cognitive behavioural intervention principles adapted to specifically address dysfunctional COVID-19 worry. The project was done in collaboration with public health authorities and made available to the public free-of-charge following testing.

#### Trials of Standard Interventions Minimally Adapted for COVID-19

The 6 trials^25-30^ that tested standard interventions minimally adapted for mental health during COVID-19 were conducted in Oman (N = 46),^25^ Malaysia (N = 61),^26^ Spain (N = 164),^27^ Iran (N = 150),^28^ Serbia (N = 104),^29^ and China (N = 104).^30^ Participants were recruited via social media,^26,29^ an internet survey,^25^ and a university email list;^27^ in 2 trials, recruitment method was not reported.^28,30^ Two trials tested standard cognitive behavioural therapy delivered individually (6 sessions)^25^ or in groups (10 sessions)^28^; both reported targeting mental health symptoms from COVID-19, but neither described COVID-19-specific intervention adaptations. Two interventions tested standard self-guided journaling (one session)^25^ or expressive writing (5 sessions)^29^ adapted by instructing participants to write about experiences during the pandemic. Two tested self-guided mindfulness apps (14 sessions)^27^ or audio recordings (5 sessions)^30^ that were described as targeting COVID-19 mental health symptoms but did not describe clear adaptations for COVID-19 challenges.

### Risk of Bias and Adequacy of Intervention Description

Risk of bias assessments are shown in Table 2 and adequacy of intervention descriptions in Table 3.

**Table 2.**
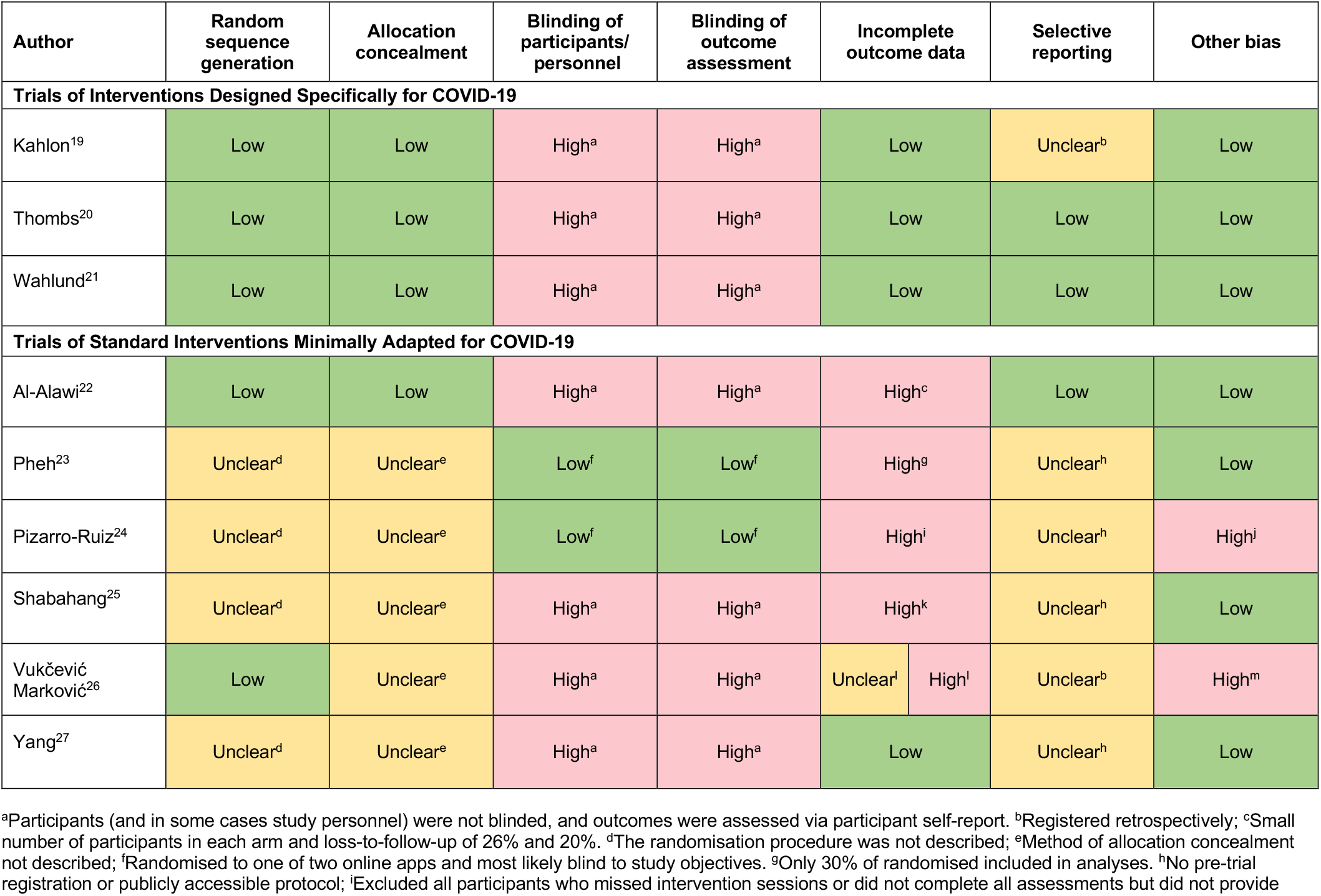
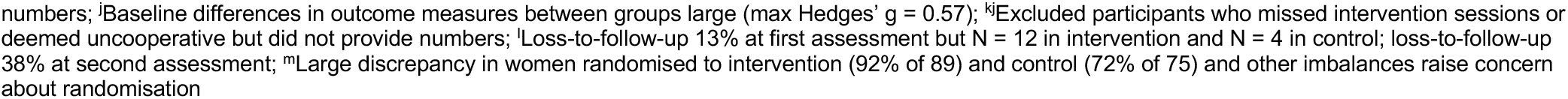
Risk of Bias of Included Trials.

| Author | Random<br>sequence<br>generation | Allocation<br>concealment | Blinding of<br>participants/<br>personnel | Blinding of<br>outcome<br>assessment | Incomplete<br>outcome data |  | Selective<br>reporting | Other bias |
| --- | --- | --- | --- | --- | --- | --- | --- | --- |
| Trials of Interventions Designed Specifically for COVID-19 |  |  |  |  |  |  |  |  |
| Kahlon<sup>19</sup> | Low | Low | High<sup>a</sup> | High<sup>a</sup> | Low |  | Unclear<sup>b</sup> | Low |
| Thombs<sup>20</sup> | Low | Low | High<sup>a</sup> | High<sup>a</sup> | Low |  | Low | Low |
| Wahlund<sup>21</sup> | Low | Low | High<sup>a</sup> | High<sup>a</sup> | Low |  | Low | Low |
| Trials of Standard Interventions Minimally Adapted for COVID-19 |  |  |  |  |  |  |  |  |
| Al-Alawi<sup>22</sup> | Low | Low | High<sup>a</sup> | High<sup>a</sup> | High<sup>c</sup> |  | Low | Low |
| Pheh<sup>23</sup> | Unclear<sup>d</sup> | Unclear<sup>e</sup> | Low<sup>f</sup> | Low<sup>f</sup> | High<sup>g</sup> |  | Unclear<sup>h</sup> | Low |
| Pizarro-Ruiz<sup>24</sup> | Unclear<sup>d</sup> | Unclear<sup>e</sup> | Low<sup>f</sup> | Low<sup>f</sup> | High<sup>i</sup> |  | Unclear<sup>h</sup> | High<sup>j</sup> |
| Shabahang<sup>25</sup> | Unclear<sup>d</sup> | Unclear<sup>e</sup> | High<sup>a</sup> | High<sup>a</sup> | High<sup>k</sup> |  | Unclear<sup>h</sup> | Low |
| Vukčević<br>Marković<sup>26</sup> | Low | Unclear<sup>e</sup> | High<sup>a</sup> | High<sup>a</sup> | Unclear<sup>l</sup> | High<sup>l</sup> | Unclear<sup>b</sup> | High<sup>m</sup> |
| Yang<sup>27</sup> | Unclear<sup>d</sup> | Unclear<sup>e</sup> | High<sup>a</sup> | High<sup>a</sup> | Low |  | Unclear<sup>h</sup> | Low |

<sup>a</sup>Participants (and in some cases study personnel) were not blinded, and outcomes were assessed via participant self-report. <sup>b</sup>Registered retrospectively; <sup>c</sup>Small number of participants in each arm and loss-to-follow-up of 26% and 20%. <sup>d</sup>The randomisation procedure was not described; <sup>e</sup>Method of allocation concealment not described; <sup>f</sup>Randomised to one of two online apps and most likely blind to study objectives. <sup>g</sup>Only 30% of randomised included in analyses. <sup>h</sup>No pre-trial registration or publicly accessible protocol; <sup>i</sup>Excluded all participants who missed intervention sessions or did not complete all assessments but did not provide
numbers; <sup>j</sup>Baseline differences in outcome measures between groups large (max Hedges' $g = 0.57$ ); <sup>kj</sup>Excluded participants who missed intervention sessions or deemed uncooperative but did not provide numbers; <sup>l</sup>Loss-to-follow-up 13% at first assessment but $N = 12$ in intervention and $N = 4$ in control; loss-to-follow-up 38% at second assessment; <sup>m</sup>Large discrepancy in women randomised to intervention (92% of 89) and control (72% of 75) and other imbalances raise concern about randomisation

**Table 3.** Reporting Quality of Interventions Based on TIDieR Checklist.

| Author<br>Dates<br>Country(ies) | Brief<br>Name | Why | Materials | Procedures | Who<br>provided | How | Where | When<br>and<br>how<br>much | Tailoring | Modification | How well<br>(Planned) | How<br>well<br>(Actual) |
| --- | --- | --- | --- | --- | --- | --- | --- | --- | --- | --- | --- | --- |
| **Trials of Interventions Designed Specifically for COVID-19** |  |  |  |  |  |  |  |  |  |  |  |  |
| Kahlon<sup>19</sup> | Yes | Yes | Partial | Yes | Partial | Yes | Yes | Yes | Yes | N/A | Yes | No |
| Thombs<sup>20</sup> | Yes | Yes | Yes | Yes | Yes | Yes | Yes | Yes | N/A | N/A | Yes | Yes |
| Wahlund<sup>21</sup> | Yes | Yes | Yes | Yes | Yes | Yes | Yes | Partial | N/A | N/A | Yes | Yes |
| **Trials of Standard Interventions Minimally Adapted for COVID-19** |  |  |  |  |  |  |  |  |  |  |  |  |
| Al-Alawi<sup>22</sup> | Yes | Partial | Partial | Yes | Partial | Yes | No | Yes | Yes | N/A | Partial | Partial |
| Pheh<sup>23</sup> | Yes | Yes | Yes | Yes | Partial | No | No | Partial | N/A | N/A | N/A | N/A |
| Pizarro-Ruiz<sup>24</sup> | Yes | Partial | Yes | Partial | No | Yes | Yes | Yes | N/A | N/A | Yes | No |
| Shabahang<sup>25</sup> | Yes | Yes | No | Partial | Yes | No | No | Yes | N/A | N/A | N/A | N/A |
| Vukčević Marković<sup>26</sup> | Yes | Yes | Yes | Yes | No | Yes | Partial | Yes | N/A | N/A | Partial | No |
| Yang<sup>27</sup> | Yes | Yes | Partial | Yes | No | Yes | Partial | Yes | N/A | N/A | N/A | N/A |

#### Trials of Interventions Designed Specifically for COVID-19

For all 3 trials,^22-24^ risk of bias was low for random sequence generation, allocation concealment, incomplete outcome data, and other bias sources. It was high for all 3 trials for blinding of participants and personnel and outcome assessment, since outcomes involved symptom self-report by non-blinded participants. Two trials^23,24^ were rated low for selective outcome reporting, because outcomes matched a priori registered outcomes; the other trial^22^ was rated unclear, because registration was retrospective. Interventions were well-described for all 3 trials with 0,^23^ 1,^24^ and 3^22^ of 12 items rated no or partial on the TIDieR Checklist.

#### Trials of Standard Interventions Minimally Adapted for COVID-19

Among the 6 trials,^25-30^ one trial25 had 3 high risk ratings, and the other 5 trials^26-30^ had between 4 and 6 unclear or high ratings out of 7 risk of bias items. Most interventions were described sub-adequately; all had 3 to 6 no or partial TIDieR Checklist ratings. Interventions either did not evaluate intervention delivery fidelity or adherence or were rated as no or partial reporting if evaluation did take place.

### Mental Health Outcomes

Intervention effects are shown in Table 4.

**Table 4.**
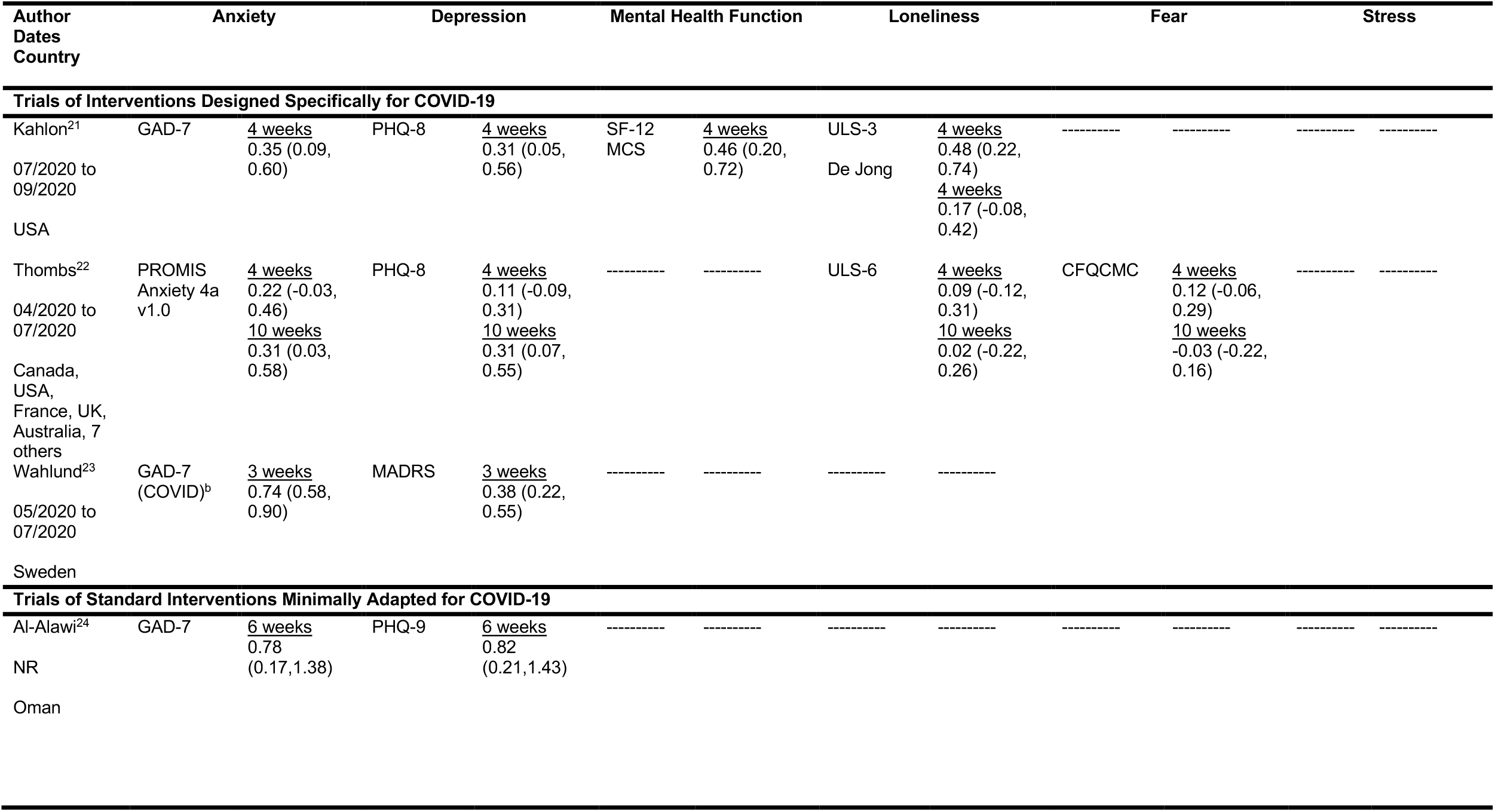
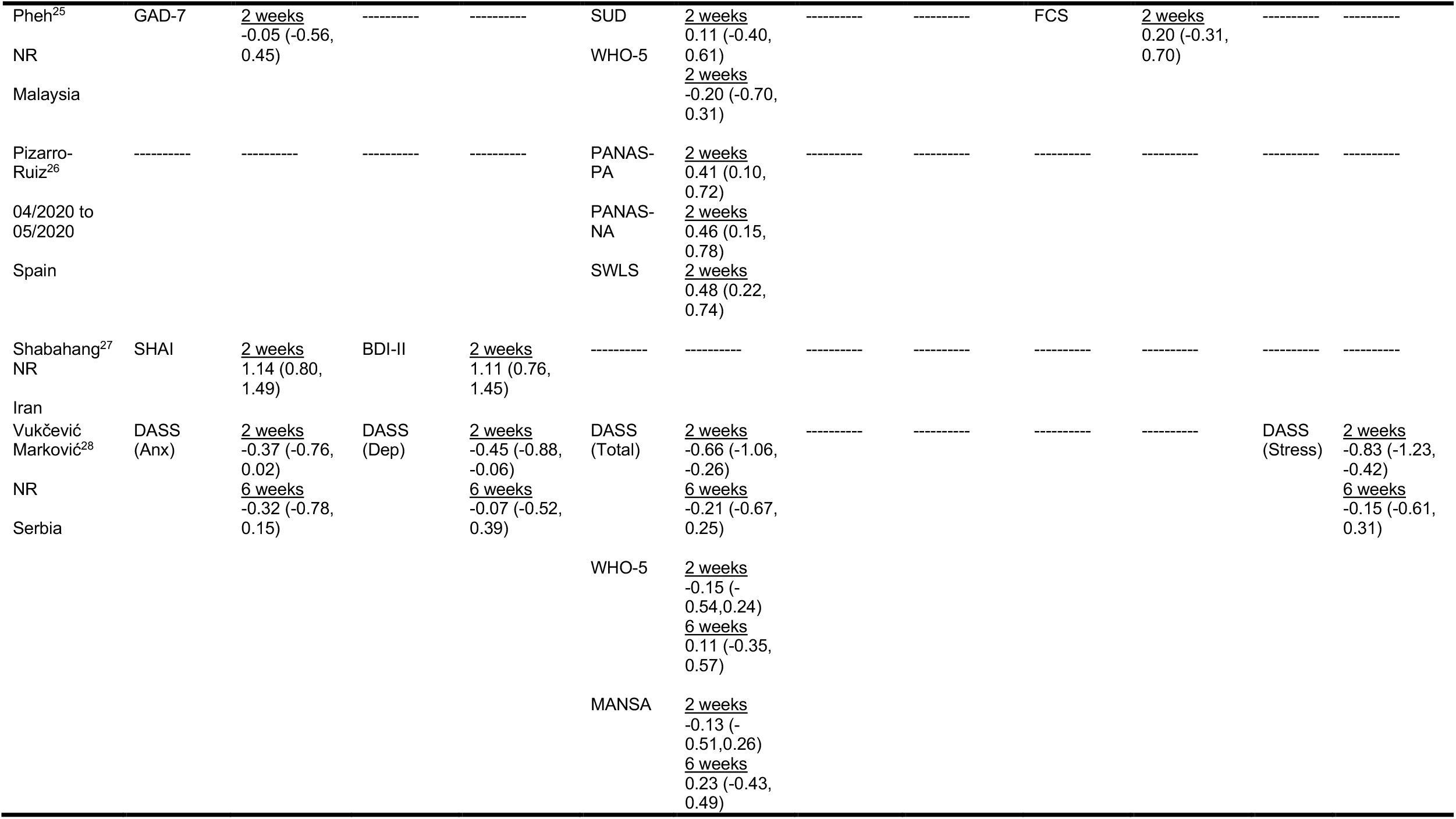
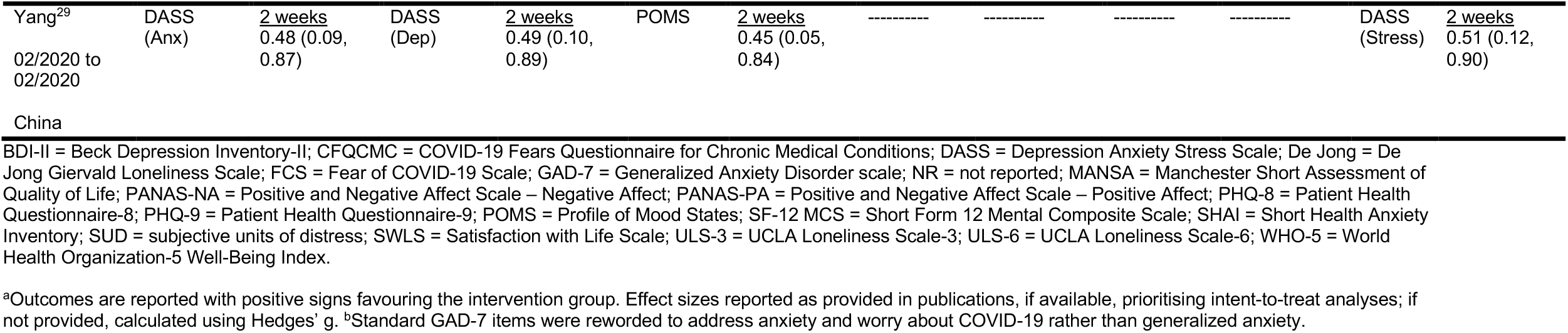
Standardized Mean Difference Effect Sizes of Mental Health Outcomes^a^.

#### Trials of Interventions Designed Specifically for COVID-19

Compared to no intervention or waitlist control, the 3 interventions^22-24^ reduced general or COVID-19-specific anxiety symptoms between SMD = 0.31 (95% CI 0.03 to 0.58)^23^ and 0.74 (95% CI 0.58 to 0.90)^24^ at the last trial assessments. Symptoms of depression were reduced by SMD = 0.31 (95% CI 0.05 to 0.56)^22^ to 0.38 (95% CI 0.22 to 0.55).^24^ For the trial in systemic sclerosis,^23^ although effects were statistically significant 10-weeks post-randomisation, they were not statistically significant immediately following the 4-week intervention. See Figures 2 and 3.

**Figure 2.**
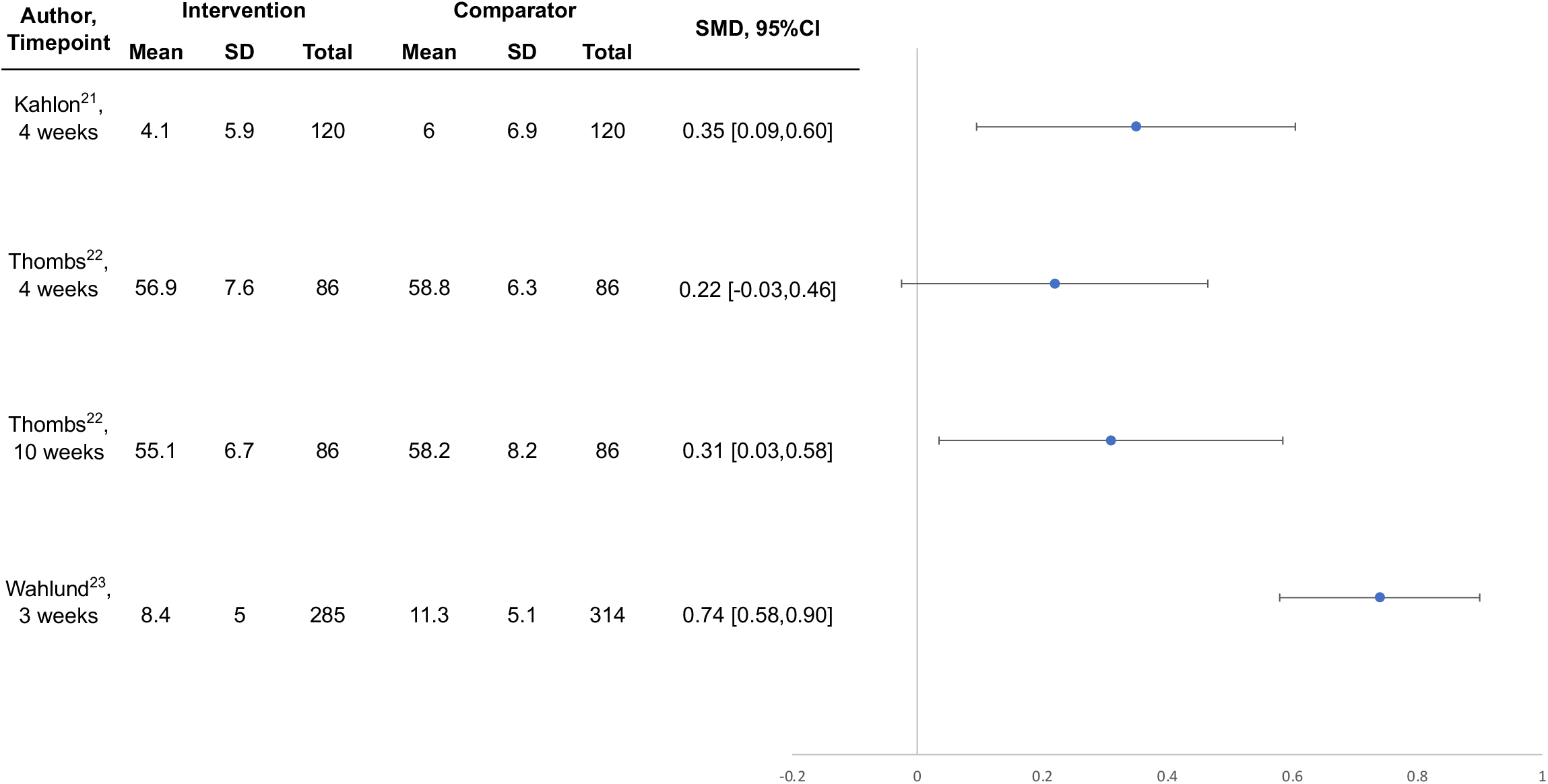
Forest Plot, Intervention Effects on Anxiety in Trials Designed Specifically for COVID-19.

**Figure 3.**
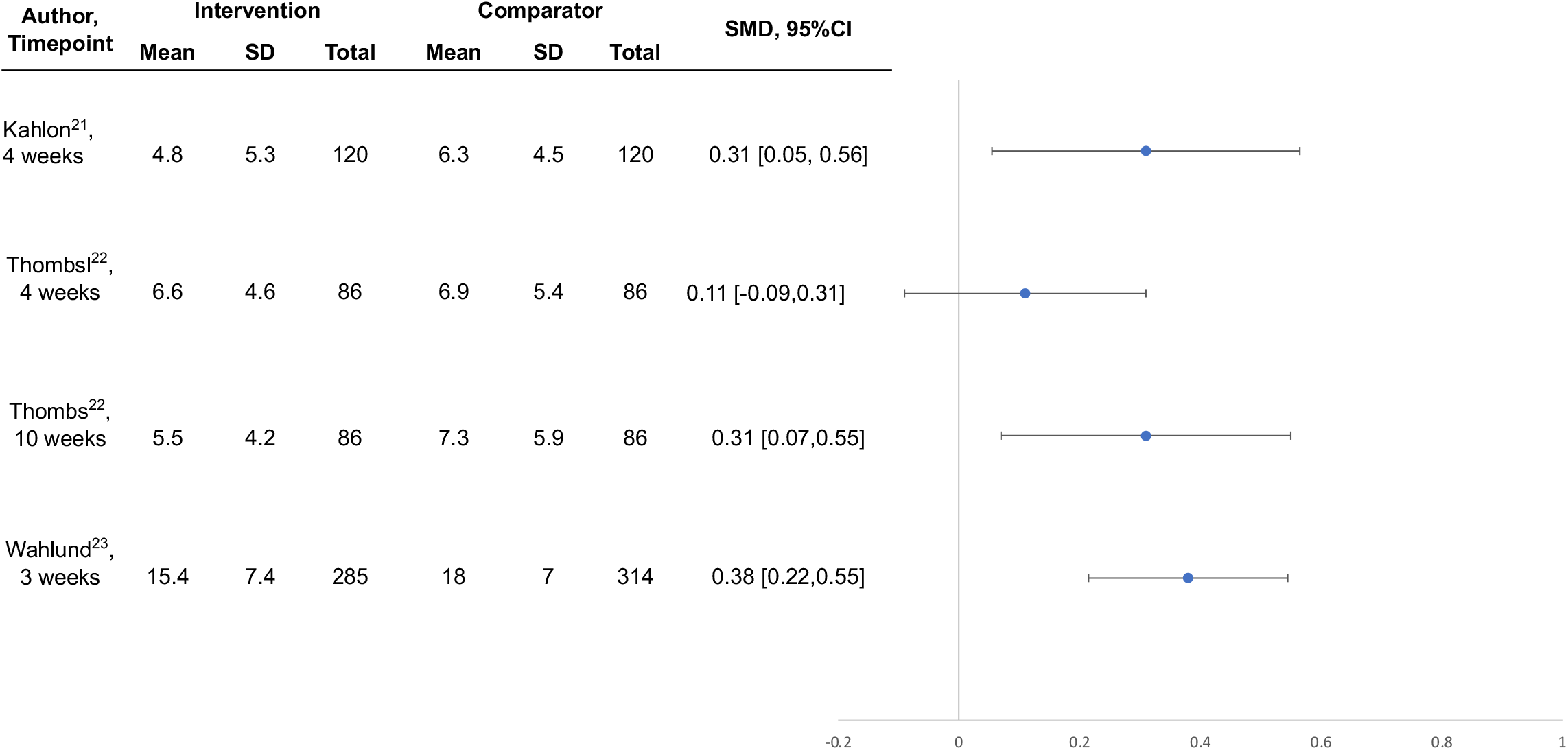
Forest Plot, Intervention Effects on Depression in Trials Designed Specifically for COVID-19.

Loneliness was reduced in the trial of lay-delivered phone calls based on one measure (SMD = 0.48, 95% CI 0.22 to 0.74) but not a second measure (SMD 0.17, 95% CI -0.08 to 0.42);^22^ neither loneliness nor fear were reduced at either assessment point in the group-based systemic sclerosis intervention.^23^ Two trials tracked adverse effects, and both reported no serious adverse effects.^23,24^

#### Trials of Standard Interventions Minimally Adapted for COVID-19

Reported effects on symptoms of anxiety and depression were between SMD = 0.78 (95% CI 0.17 to 1.38) and 1.14 (95% CI 0.80 to 1.49) for the individual and group cognitive behavioural therapy interventions.^25,28^ Effects for single-session mindfulness-based journaling were close to null and not statistically significant.^26^ For 5 sessions of expressive writing, of 12 outcome assessments, none favoured the intervention, but 3 were statistically significant and large in favour of the no-intervention control.^29^ The two studies that tested app-based or audio-recorded mindfulness interventions reported statistically significant effects on several variables in favour of the intervention.^27,30^ The only trial that tracked adverse effects reported no adverse effects.^24^

## DISCUSSION

Scalable, feasibly delivered interventions are needed to address community mental health implications of COVID-19 that will likely persist beyond the pandemic. We identified 3 well-conducted trials of potentially scalable interventions designed to address COVID-19 mental health in the general public24 and among people vulnerable in COVID-19 due to age and pre-existing medical conditions.^22,23^ A self-guided online intervention that targeted COVID-19-specific dysfunctional worry reduced COVID-19 anxiety by SMD = 0.74 (95% CI 0.58 to 0.90) and depression symptoms by SMD = 0.38 (95% CI 0.22 to 0.55) in the Swedish general public.^24^ A lay-delivered supportive telephone intervention reduced anxiety and depression symptoms and improved mental health function by SMD = 0.31 (95% CI 0.05 to 0.56) to SMD =

0.46 (95% CI 0.20 to 0.72) among homebound older adults in the United States.^22^ A multifaceted group-based intervention for people with systemic sclerosis from 12 countries, which included peer-led support plus professionally delivered mental health coping strategies, did not significantly reduce mental health outcomes immediately post-intervention, but anxiety (SMD = 0.31; 95% CI 0.03 to 0.58) and depression (SMD = 0.31; 95% CI 0.07 to 0.55) symptoms significantly improved 6 weeks later.^23^ These effect sizes are comparable to effects from treating major depressive disorder with antidepressants (SMD = 0.31)^31^ or from cognitive behavioural therapy for depression in primary care (SMD = 0.22);^32^ both considered standard health care. We did not identify any trials of interventions for children or adolescents.

We identified 6 trials that tested delivery of standard psychological interventions without significant adaptation for COVID-19, including individual or group-based cognitive behavioural therapy,^25,28^ expressive writing,^26,29^ and self-guided mindfulness apps or audio recordings.^27,30^ There were serious concerns, however, about risk of bias and adequacy of reporting in all of these trials, which reduced confidence in results.

Governments and health care providers around the world need effective, scalable interventions to meet the challenges of population mental health in COVID-19, including digital33 and other types of interventions.^9,10^ Our findings show that digital interventions for the general public and lay-delivered or peer-supported telephone or videoconference interventions for people who are vulnerable due to age or pre-existing medical conditions may be effective solutions.

The finding that a self-guided internet intervention reduced both anxiety and depression symptoms is consistent with a growing body of evidence that internet-based psychological interventions may be an effective first-line strategy for many people. They are likely not as effective as in-person or guided internet-based therapies and may not be appropriate for people with severe or unremitting illness.^34^ However, consistent with the findings of the study by Wahlund et al.^24^ in the present review, some studies have found that estimates of effectiveness approach those of guided formats, including for anxiety and depressive disorders.^34,35^

Evidence is mixed on the effectiveness of “befriending” or social support-based interventions delivered via video-based communication, online discussion groups and forums, or telephone.^36^ However, factors that appear to be associated with greater likelihood of effectiveness include shared experiences or characteristics among participants and the ability of participants to speak freely and develop relationships.^36^ These were key components of the two trials that used an empathetic telephone calling strategy22 and peer-moderated videoconference-based groups for people with the rare autoimmune disease systemic sclerosis.^23^

We did not identify any trials of interventions for children or adolescents, and it is not known to what degree self-guided or lay-and peer-support interventions would be effective. Unfortunately, as of April 29, 2021, no trials that planned to test mental health interventions with children or adolescents in COVID-19 had been registered.^4^

Strengths of our systematic review include using rigorous best-practice methods; searching 9 databases, including 2 Chinese databases; not restricting inclusion by language; and the ability to update rapidly as evidence emerges via our living systematic review approach. There are also limitations. First, we identified only three trials designed specifically to address COVID-19 mental health challenges. Second, the quality and plausibility of results of many trials we encountered was concerning. We were not able to verify the accuracy of what was reported in many trials and thus only described results from those trials in appendices. Third, we are not able to rule out the possibility that publication bias, or even censorship,^37^ may have influenced our results. Fourth, the evidence base is rapidly evolving, and main results could change, although our living systematic review format will allow rapid updating as this occurs.

In summary, we identified 3 trials of interventions designed specifically to meet the needs of the general public or vulnerable populations in COVID-19. Together, they suggest that self-guided online interventions targeted to challenges faced by the public in COVID-19 can effectively support mental health and that lay-or peer-delivered interventions may be an effective strategy for vulnerable populations. Additional trials are needed, particularly to address mental health challenges among children and adolescents and among diverse populations, both currently and as pandemic conditions reside.

## Data Availability

All data from the living systematic review are available in the present manuscript and its supplementary material or online

https://www.depressd.ca/research-question-3-intervention

## Author Contributions

YS, DBR, AB, and BDT were responsible for the study conception and design. JTB was responsible for the design of the database searches. AK carried out the searches. OB, YWang, KL, XJ, AK, CH, YS, YWu, SM, DBR, ITV, AT, TDS, AY, MA, and BDT contributed to data extraction, coding, evaluation of included studies. OB, CH, and YS were responsible for study coordination. OB, AB, and BDT were involved in data analysis, and all authors were involved in interpretation of results. OB and BDT drafted the manuscript. All authors provided a critical review and approved the final manuscript. OB, CH, YS and BDT had full access to all the data in the study and take responsibility for the integrity of the data and the accuracy of the data analyses. BDT is the corresponding author and attests that all listed authors meet authorship criteria and that no others meeting the criteria have been omitted.

## Data Sharing

All data from the living systematic review are available in the present manuscript and its supplementary material or online (https://www.depressd.ca/research-question-3-intervention).

## Acknowledgements

The living systematic review was funded by the Canadian Institutes of Health Research (CMS-171703; MS1-173070) and McGill Interdisciplinary Initiative in Infection and Immunity Emergency COVID-19 Research Fund (R2-42). YWu was supported by a Fonds de recherche du Québec – Santé (FRQS) Postdoctoral Training Fellowship. DBR was supported by a Vanier Canada Graduate Scholarship. AB was supported by FRQS senior researcher salary awards. BDT was supported by a Tier 1 Canada Research Chair. No sponsor or funder was involved in the study design; in the collection, analysis and interpretation of the data; in the writing of the report; or in the decision to submit the paper for publication.

## FIGURE LEGENDS

**Figure 1.** PRISMA Flow Diagram

**Figure 2.** Forest plot of effects on symptoms of anxiety among interventions designed to address COVID-19 mental health challenges

**Figure 3.** Forest plot of effects on symptoms of depression among interventions designed to address COVID-19 mental health challenges

