## Appendices for "Effects of COVID-19 Mental Health Interventions among Community-based Children, Adolescents, and Adults: A Living Systematic Review of Randomised Controlled Trials"

**Appendix 1: Search Strategies**

Search strategies can be found in the project folder on the Open Science Framework:

<https://osf.io/96csg/>

**Appendix 2. Inclusion and Exclusion Coding Guides for Main Interventions Review Plus Additional Criteria for Present Report**

Title and Abstract Review:

**Exclude: not original human data or a case study or case series.** If it is clear from the title and abstract that the article is not an original report of primary data, but, for example, a letter, editorial, systematic review or meta-analysis, or it is a single case study or case series, then it is excluded. Studies reporting only on animal, cellular, or genetic data are also excluded. Conference abstracts are included.

**Exclude: not a study of any population affected by the COVID-19 outbreak.** Eligible studies must be initiated after China's first announcement to the WHO on December 31, 2019. If it is clear from the title or abstract that the study is not about any population affected by the COVID-19 outbreak, it is excluded. Studies that include fewer than 10 subjects, are excluded.

**Exclude: intervention does not target mental health.** If it is clear from the title or abstract that the study is not about an intervention or is an intervention, but the intervention does not primarily target mental health, then it will be excluded. Mental health must be the primary trial outcome if a primary outcome or outcomes are stated.

**Exclude: not a randomized or non-randomized controlled trial (RCT) with eligible comparators:** If it is clear from the title or abstract that the study is not a RCT or non-randomized controlled trial that compares an intervention designed to improve any aspect of mental health during the COVID-19 pandemic to (1) any inactive control condition (e.g., no treatment, waitlist control) or to (2) another eligible intervention designed to mental health, then it will be excluded.

**Include: study eligible to be included in full-text review.**

Full-text Review:

**Exclude: not original human data or a case study or case series.** If the article is not an original report of primary data, but, for example, a letter, editorial, systematic review or meta-analysis, or it is a single case study or case series, then it is excluded. Studies reporting only on animal, cellular, or genetic data are also excluded. Conference abstracts are included.

**Exclude: not a study of any population affected by the COVID-19 outbreak.** Eligible studies must be initiated after China's first announcement to the WHO on December 31, 2019. If the study is not about any population affected by the COVID-19 outbreak, it is excluded. Studies that include fewer than 10 subjects, are excluded.

**Exclude: intervention does not target mental health.** If the study is not about an intervention or is an intervention, but the intervention does not primarily target mental health, then it will be excluded. Mental health must be the primary trial outcome if a primary outcome or outcomes are stated.

**Exclude: not a randomized or non-randomized controlled trial (RCT) with eligible comparators:** If the study is not a RCT or non-randomized controlled trial that compares an intervention designed to improve any aspect of mental health during the COVID-19 pandemic to (1) any inactive control condition (e.g., no treatment, waitlist control) or to (2) another eligible intervention designed to mental health, then it will be excluded.

**Include: study eligible for inclusion in systematic review.**

**Additional Criteria for Present Report: (1) randomised controlled trial; (2) assessed outcomes at least one week after intervention initiation; (3) population not hospitalized or quarantined due to COVID-19; (4) trial report accuracy verified by authors.**

**Appendix 3. Otherwise Eligible Unverified Trials (Authors did not respond to requests for verification)**

**Information also available at https://www.depressd.ca/research-question-3-intervention**

***Trial Characteristics (N = 13)***

| Study Details | Participants | Country | Intervention | Comparator | Dates of Data Collection | N Intervention (Analyzed) | N Comparator (Analyzed) | Outcome Domain(s) | Mean (SD) Age | % Female or Women |
| --- | --- | --- | --- | --- | --- | --- | --- | --- | --- | --- |
| Authors did not respond to requests for verification (N= 7) | | | | | | | | | | |
| Li, Acta Psychologica Sinica, 2020  https://dx.doi.org/10.3724/SP.J.1041.2020.00886 | Chinese adults from various provinces | China | Participants completed a value selection scale and wrote about a value they chose as most important | Participants completed a value selection scale and wrote about a value they chose as least important | 02/2020-02/2020 | 96 | 91 | Depression, Anxiety | 24 (7) | 63 |
| Cui,  China Journal of Health Psychology,  2020  https://kns.cnki.net/kcms/detai l/11.5257.R.20200828.1458.019.html | Adults over 18 who can use smartphone | China | 2-week, 2x per week online mindfulness-based stress-reduction group therapy | Online mental health education | 02/2020 | 74 | 74 | Mental Health Function,  Depression,  Anxiety | Intervention  36 (8)    Comparator  37 (9) | 61 |
| [Fan,  Journal of Environmental and Occupational Medicine,  2020   https://doi.org/10.13213/j.cnki.jeom.2020.20130](https://doi.org/10.13213/j.cnki.jeom.2020.20130) | Volunteer undergraduate students with anxiety symptoms from a university in Hangzhou | China | 7-day Wechat group communication plus solution-focused intervention related questions, which are positively evaluated and answered by researchers in private messages 15min a day | Wechat group communication for 7 days, including researchers encouraging participants and participants expressing their emotions | 15/02/2020 - 21/02/2020 | 71 | 71 | Mental Health Function,  Anxiety | NR | 49 |
| Preuss, Internet Interventions  2021  <https://doi.org/10.1016/j.invent.2021.100388> | Parents: German adults taking care of at least one child aged 3 to 18 years who had attended a kindergarten or school prior to the pandemic outbreak | Germany and Austria | Self-compassion intervention, including a 20min video covering psychoeducation and guided self compassion practice. Booster session included a 10min guided video session. Three email reminders aimed to support the daily practice were sent | Waitlist control | 04/2020-06/2020 | 75 | 69 | Stress | 41 (6) | 93 |
| Dincer, Explore, 2020  https://doi.org/10.1016/j.explore.2020.11.012 | Nurses who care for patients infected with COVID-19 | Turkey | A single 20-minute online EET (Emotional Freedom Techniques) session | Participants were asked to stay comfortable in a calm and tranquil environment for the next 15 minutes | 05/2020-05/2020 | 35 | 37 | Mental Health Function, Anxiety, Burnout | 34 (10) | 89 |
| [Deng,  Journal of Nursing (China), 2020  https://doi.org/10.16460/j.issn1008-9969.2020.21.064](https://doi.org/10.16460/j.issn1008-9969.2020.21.064) | Nurses on the frontline against COVID-19 for greater or equal to 1 week and are in healthy condition | China | Internet-based unstructured group counselling in groups of 10 or 12 delivered 1x per 2 days in 10 days from 19:00 to 21:05, each member of the group has 10min to discuss the topic, with a time allocation of "4min+4min+2min", where the first 4min is for sharing personal experience, the second for experience communication, and the rest 2min for leaders of the group to make a summary | Standard tutorial on how to prevent and control COVID-19 and the related psychology knowledge | NR | 42 | 43 | Mental Health Function, Stress | 20-30: 36%;  31-40: 29%;  41-50: 34% | NR |
| [Song, World Chinese Journal of Digestology, 2021  https://dx.doi.org/10.11569/wcjd.v29.i1.48](https://dx.doi.org/10.11569/wcjd.v29.i1.48) | Elderly patients with gastroesophageal reflux disease and without severe organic diseases in digestive system; no severe psychiatric or neurological disorder, or with severe multiple organ function deficiency, or malignant tumor carrier, or with history of abdominal operation | China | Routine medical care plus 30min mindfulness-based stress reduction therapy once per two days for 8 times, content including mindfulness breathing exercise, body scan meditation, and expression of the self in relation to COVID-19 | Routine medical care, including diet care, health education, medication care, sports care, psychological care, and disease-related knowledge consultation. | NR | 60 | 60 | Depression,  Anxiety, Stress,  Mental Health Function | Intervention 71 (9)    Comparator 69 (10) | 47 |
| Corresponding author contact information was not available (N=6) | | | | | | | | | | |
| [Li,  Jiangxi Medical Journal,  2020   https://www.doi.org/10.3969 /j.issn.1006-2238.2020.07.009](https://www.doi.org/10.3969/j.issn.1006-2238.2020.07.009) | Voluntary participants struggling with emotional and sleep disturbance during COVID-19 | China | Group Mindfulness-based stress reduction intervention delivered online 2x per week, each session lasts 1.5h, the first week's content including introducing and familiarizing participants with the intervention, the second week focuses on body scan meditation, and the third week focuses on 3min-breathing exercise | Online group supportive psychotherapy 2x per week in 3 weeks, with each session lasts 1.5h, focusing on adaptation strategies and positive mindset | NR | 36 | 36 | Anxiety | 28 (9) | 47 |
| [Zhang, Henan Medical Research, 2020  https://www.doi.org/10.3969%20/j .issn.1004-437X.2020.32.001](https://www.doi.org/10.3969%20/j.issn.1004-437X.2020.32.001) | First-line support nurses with no mental or psychological disorder history or experiencing recent major life events | China | WeChat group plus meditation training 30min per session, 1x per day, 5x per week for 4 weeks, including muscle relaxation, breathing exercise, emotional awareness training. | WeChat group exchange of knowledge with regard to psychological wellbeing | 03/2020-04/2020 | 29 | 25 | Anxiety | Intervention 30 (4)  Comparator 32 (3) | NR |
| Li, Health Guide, 2020  https://d.wanfangdata.com.cn/periodical/ChlQZXJpb2RpY2FsQ0hJTmV3UzIwMjEwNDE1EhF5c2Jqem4teDIwMjA1MTI3ORoIbzdvZWRzMWM%3D | Pregnant women with gestational weeks 28-31+6 and had oral glucose tolerance test at 24-28 weeks; no sign of threatened premature labor or pregnancy complications; and had degree greater than or equal to high school, with no internal diseases or psychiatric disorders | China | Health education and psychological guidance via WeChat platform, where questions posed by participants were answered by doctors; self-monitored blood glucose level reported by participants was instructed by nursing staff; 30min weekly nutrition and COVID-19 education session; and relaxation techniques provided by counselling psychologists, participants with anxiety and depression symptoms were targeted and provided with 2 group therapies | Screened twice 2 weeks and 4 weeks after the study initiation and checked for blood glucose level; online psychological counselling recommended for participants with anxiety symptoms | NR | 72 | 72 | Depression, Anxiety | 29 (3) | 100 |
| [Li,  Today Nurse 2020  https://www.doi.org/10.19792%2 0/j.cnki.1006-6411.2020.35.038](https://www.doi.org/10.19792%20/j.cnki.1006-6411.2020.35.038) | Children 8 years or older undergoing nebulizer therapy for at least 1 week and their parents | China | Psychological intervention, including educating about nebulizer therapy information and COVID-19 related knowledge to alleviate anxiety and achieve enhanced self-protection | Standard nursing, including informing nebulizer therapy instructions | NR | 30 | 30 | Anxiety | Intervention Children: 11 (3) Parents: 37 (8)  Comparator Children: 11 (2) Parents: 38 (10) | 72 |
| Wei, Chinese Community Doctors, 2021  10.3969/j.issn.1007-614x.2021.07.091 | Adult community residents with >2 years of residence | China | Application of "Internet+" model in health management, the health education content was delivered daily at 9am via WeChat group | Health management via telephone, with the content focusing on COVID-related information, including self-preventative and self-protection strategies | NR | 140 | 140 | Depression, Anxiety, Mental health function | Intervention 42 (6)  Comparator 42 (7) | 44 |
| Huang, Journal of Chengdu Medical College, 2020  https://kns.cnki.net/kcms/detail/detail.aspx?dbcode=CAPJ&dbname=CAPJLAST&filename=CDYU20200903001&v=cH4TTge1SJ%25mmd2F%25mmd2BGo6kUslNdxX4ZCkL0rg0bexMC4MuBpK7eeZH3WbOaB%25mmd2FghEJeas1L | Medical staff working at isolation ward for greater or equal to one week | China | Mental health education intervention delivered via videos from 8:00-8:40pm everyday for 8 weeks, and informative crisis handling brochure | 40min daily mindfulness-based stress reduction intervention for 8 weeks, where the mindfulness decompressing training theme varies weekly and each lasts 5 days | NR | 60 | 58 | Stress, Anxiety, Depression, Mental health function | Intervention 33 (7)  Comparator 32 (5) | 77 |

***Effect Sizes of Mental Health Outcomes (positive effects favour intervention group)***

| Study Details | | Outcome Domain | | Outcome Measure | | Primary/ Secondary | Raw Mean Difference (95% CI) Effect Size | | | Hedges' g (95% CI) Effect Size |
| --- | --- | --- | --- | --- | --- | --- | --- | --- | --- | --- |
| Authors did not respond to requests for verification (N= 7) | | | | | | | | | | |
| Li, Acta Psychologica Sinica, 2020 | | Depression | | BDI | | NR | 0.02 (-1.88, 1.92) | | | 0.00 (-0.28, 0.29) |
| Li, Acta Psychologica Sinica, 2020 | | Anxiety | | ZSAS | | NR | 2.18 (0.35, 4.01) | | | 0.34 (0.05, 0.63) |
| Cui, China Journal of Health Psychology,  2020 | | Mental Health Function | | SSS-8 | | NR | 7.78 (5.44, 10.12) | | | 1.07 (0.73, 1.42) |
| Cui, China Journal of Health Psychology,  2020 | | Depression | | PHQ-9 | | NR | 3.13 (1.85, 4.41) | | | 0.79 (0.45, 1.13) |
| Cui, China Journal of Health Psychology,  2020 | | Anxiety | | GAD-7 | | NR | 2.02 (1.02, 3.02) | | | 0.65 (0.32, 0.98) |
| Fan, Journal of Environmental and Occupational Medicine, 2020 | | Mental Health Function | | PANAS (PA) | | NR | 3.43 (1.68, 5.18) | | | 0.65 (0.31, 0.99) |
| Fan, Journal of Environmental and Occupational Medicine, 2020 | | Mental Health Function | | PANAS (NA) | | NR | 3.73 (2.69, 4.77) | | | 1.19 (0.83, 1.55) |
| Fan, Journal of Environmental and Occupational Medicine, 2020 | | Anxiety | | ZSAS | | NR | 8.95 (6.71, 11.19) | | | 1.32 (0.95, 1.68) |
| Preuss, Internet Interventions  2021 | | Stress  (Cognitive reappraisal intervention) | | PSS-10 | | Primary | 4.59 (2.18, 7.00) | | | 0.63 (0.29, 0.97) |
| Preuss, Internet Interventions  2021 | | Stress  (Cognitive reappraisal intervention) | | PSQ-PS (global) | | Primary | 3.85 (0.50, 7.20) | | | 0.38 (0.05, 0.71) |
| Preuss, Internet Interventions  2021 | | Stress  (Self-compassion intervention) | | PSS-10 | | Primary | 3.12 (0.82, 5.42) | | | 0.44 (0.11, 0.78) |
| Preuss, Internet Interventions  2021 | | Stress  (Self-compassion intervention) | | PSQ-PS (global) | | Primary | 2.11 (-1.26, 5.48) | | | 0.21 (-0.12, 0.53) |
| Dincer, Explore, 2020 | | Mental Health Function | | SUD | | NR | 4.55 (3.90, 5.20) | | | 3.25 (2.54, 3.96) |
| Dincer, Explore, 2020 | | Anxiety | | STAI | | NR | 32.18 (29.17, 35.19) | | | 4.98 (4.03, 5.92) |
| Dincer, Explore, 2020 | | Burnout | | BS | | NR | 0.95 (0.52, 1.38) | | | 1.02 (0.53, 1.52) |
| [Deng, Journal of Nursing (China), 2020](https://doi.org/10.16460/j.issn1008-9969.2020.21.064) | | Mental Health Function | | PANAS-NA | | NR | 13.35 (12.23, 14.47) | | | 5.09 (4.21, 5.98) |
| Deng, Journal of Nursing (China), 2020 | | Stress | | PSS | | NR | 7.93 (6.59, 9.27) | | | 2.54 (1.96, 3.11) |
| Song, World Chinese Journal of Digestology, 2021 | | Depression | | DASS-21 | | NR | 0.72 (0.36, 1.08) | | | 0.72 (0.35, 1.09) |
| Song, World Chinese Journal of Digestology, 2021 | | Anxiety | | DASS-21 | | NR | 0.38 (0.02, 0.74) | | | 0.38 (0.02, 0.74) |
| Song, World Chinese Journal of Digestology, 2021 | | Stress | | DASS-21 | | NR | 0.64 (0.26, 1.02) | | | 0.61 (0.24, 0.98) |
| Song, World Chinese Journal of Digestology, 2021 | | Mental Health Function | | POMS | | NR | -0.13 (-4.52, 4.26) | | | -0.01 (-0.37, 0.35) |
| Corresponding author contact information was not available (N=6) | | | | | | | | | | |
| Li, Jiangxi Medical Journal, 2020 | Anxiety | | ZSAS | | NR | | | 14.46 (10.94, 17.98) | 1.91 (1.35, 2.47) | |
| Zhang, Henan Medical Research, 2020 | Anxiety | | ZSAS | | NR | | | Median (Q1, Q3): 32.0 (29.0, 34.5) | Not enough information to calculate | |
| Li, Health Guide, 2020 | Anxiety | | HAM-A | | NR | | | NR | Not enough information to calculate | |
| Li, Health Guide, 2020 | Depression | | ZSDS | | NR | | | NR | Not enough information to calculate | |
| Li, Today Nurse 2020 | Anxiety (Children) | | SCAS | | NR | | | 11.96 (0.29, 23.63) | 0.52 (0.00, 1.04) | |
| Li, Today Nurse 2020 | Anxiety (Parents) | | ZSAS | | NR | | | 8.61 (0.34, 16.88) | 0.53 (0.01, 1.05) | |
| Wei, Chinese Community Doctors, 2021 | Anxiety | | ZSAS | | NR | | | 2.15 (1.76, 2.54) | 1.30 (1.04, 1.56) | |
| Wei, Chinese Community Doctors, 2021 | Depression | | ZSDS | | NR | | | 2.41 (2.12, 2.70) | 1.94 (1.66, 2.22) | |
| Wei, Chinese Community Doctors, 2021 | Mental Health Function | | SF-36 | | NR | | | 5.48 (3.80, 7.16) | 0.77 (0.52, 1.01) | |
| Huang, Journal of Chengdu Medical College, 2020 | Depression | | PHQ-9 | | NR | | | NR | Not enough information to calculate | |
| Huang, Journal of Chengdu Medical College, 2020 | Anxiety | | GAD-7 | | NR | | | NR | Not enough information to calculate | |

BDI= Beck Depression Inventory; BS= Burnout Scale; DASS-21= Depression, Anxiety and Stress Scale; GAD-7= Generalized Anxiety Disorder; HAM-A= Hamilton Anxiety Rating Scale; NR= Not Reported; PANAS= Positive and Negative Affect Schedule; PHQ-9= Patient Health Questionnaire; POMS= Mood State Scale; PSQ-PS= Parental Stress Questionnaire- Parental Stress; PSS-10= Perceived Stress Scale; SCAS= Spence Children's Anxiety Scale; SF-36= Short-Form-36 (SF-36) Health Survey - Mental Health Component; SSS-8= Somatic Symptom Scale; STAI= State-Trait Anxiety Inventory; SUD= Subjective Units of Distress Scale; ZSAS= Zung Self-rating Anxiety Scale; ZSDS= Zung Self-rating Depression Scale

***Risk of Bias***

| **Study Details** | **Random sequence generation** | **Allocation concealment** | **Blinding of participants/personnel** | **Blinding outcome assessment** | **Incomplete outcome data** | **Selective reporting** | **Free of other bias** |
| --- | --- | --- | --- | --- | --- | --- | --- |
| **Authors did not respond to requests for verification (N= 7)** | | | | | | | |
| Li, Acta Psychologica Sinica, 2020 | Unclear risk^a^ | Unclear risk^a^ | Low risk | Low risk | Unclear risk^b^ | Unclear risk^c^ | Low risk |
| Cui,  China Journal of Health Psychology,  2020 | Low risk | Unclear risk^d^ | High risk | High risk | Unclear risk^e^ | Unclear risk^c^ | Low risk |
| Fan,  Journal of Environmental and Occupational Medicine,  2020 | Unclear risk^f^ | Unclear risk^d^ | High risk | Unclear risk^g^ | Low risk | Unclear risk^c^ | Low risk |
| Preuss, Internet Interventions  2021 | Low risk | Low risk | High risk | High risk | High risk | Low risk | Low risk |
| Dincer, Explore, 2020 | Low risk | Low risk | High risk | High risk | Unclear risk^h^ | Unclear risk^c^ | Low risk |
| Deng,  Journal of Nursing (China), 2020 | Low risk | Unclear risk^d^ | High risk | High risk | Low risk | Unclear risk^c^ | Low risk |
| [Song, World Chinese Journal of Digestology, 2021](https://dx.doi.org/10.11569/wcjd.v29.i1.48) | Unclear risk^a^ | Unclear risk^d^ | High risk | High risk | Unclear risk^e^ | Unclear risk^c^ | Low risk |
| **Corresponding author contact information was not available (N=6)** | | | | | | | |
| [Li, Jiangxi Medical Journal, 2020](https://www.doi.org/10.3969/j.issn.1006-2238.2020.07.009) | Low risk | Unclear risk^i^ | Unclear risk^j^ | Unclear risk^g^ | Low risk | Unclear risk^c^ | Low risk |
| [Zhang, Henan Medical Research, 2020](https://www.doi.org/10.3969%20/j.issn.1004-437X.2020.32.001) | Low risk | Unclear risk^i^ | High risk | High risk | Low risk | Unclear risk^c^ | Low risk |
| Li, Yang Sheng Bao Jian, 2020 | Low risk | Unclear risk^i^ | High risk | High risk | Low risk | Unclear risk^c^ | Low risk |
| Li, Today Nurse 2020 | Low risk | Unclear risk^i^ | Unclear risk^j^ | Unclear risk^g^ | Low risk | Unclear risk^c^ | Low risk |
| Wei, Chinese Community Doctors, 2021 | Unclear risk^f^ | Unclear risk^i^ | High risk | High risk | Low risk | Unclear risk^c^ | Low risk |
| Huang, Journal of Chengdu Medical College, 2020 | Unclear risk^f^ | Unclear risk^i^ | High risk | High risk | Low risk | Unclear risk^c^ | Low risk |

^a^The authors describe sequence generation and allocation but it's unclear whether participant numbers were assigned randomly; ^b^The reason for attrition is unclear; ^c^A protocol or registration with pre-specified outcomes was not provided; ^d^The allocation procedure isn't described; ^e^The proportion of randomized participants who dropped out and the method used to account for missing data is unclear; ^f^The randomization procedure is not described; ^g^Outcomes were participant reported and it's unclear whether participants were blinded to their allocation; ^h^The reason for attrition and the method used to account for missing data is unclear; ^i^The author did not provide enough information to permit judgement; ^j^It is unclear whether participants and personnel were blinded;

***Intervention Reporting***

| **Study Details** | **Brief Name** | **Why** | **Materials** | **Procedures** | **Who provided** | **How** | **Where** | **When and how much** | **Tailoring** | **Modification** | **How well (Planned)** | **How well (Actual)** |
| --- | --- | --- | --- | --- | --- | --- | --- | --- | --- | --- | --- | --- |
| **Authors did not respond to requests for verification (N= 7)** | | | | | | | | | | | | |
| Li, Acta Psychologica Sinica, 2020 | No | Yes | No | Partial^a^ | No | No | No | Partial^b^ | N/A | N/A | N/A | N/A |
| [Cui,  China Journal of Health Psychology,  2020](https://kns.cnki.net/kcms/detail/11.5257.R.20200828.1458.019.html) | Yes | Yes | Partial^c^ | Yes | Partial^d^ | Yes | Partial^e^ | Yes | N/A | N/A | N/A | N/A |
| [Fan,  Journal of Environmental and Occupational Medicine,  2020](https://doi.org/10.13213/j.cnki.jeom.2020.20130) | Yes | Yes | Yes | Yes | Partial^d^ | Yes | Yes | Yes | N/A | N/A | N/A | N/A |
| [Preuss, Internet Interventions](https://www.doi.org/10.31234/osf.io/7fue9)  [2021](https://www.doi.org/10.31234/osf.io/7fue9) | Yes | Yes | No | Yes | No | Yes | Partial^f^ | Yes | Yes | N/A | Yes | Yes |
| [Dincer, Explore, 2020](https://doi.org/10.1016/j.explore.2020.11.012) | Yes | Yes | Yes | Yes | Yes | Yes | Partial^g^ | Yes | Yes | N/A | N/A | N/A |
| [Deng,  Journal of Nursing (China), 2020](https://doi.org/10.16460/j.issn1008-9969.2020.21.064) | Yes | Yes | No | Yes | Yes | Yes | Yes | Yes | N/A | N/A | N/A | N/A |
| [Song, World Chinese Journal of Digestology, 2021](https://dx.doi.org/10.11569/wcjd.v29.i1.48) | Yes | Yes | Partial^c^ | Partial^h^ | No | Yes | Yes | Yes | N/A | N/A | N/A | N/A |
| **Corresponding author contact information was not available (N=6)** | | | | | | | | | | | | |
| Li, Jiangxi Medical Journal, 2020 | Yes | Yes | No | Yes | No | Yes | Partial^j^ | Yes | N/A | N/A | N/A | N/A |
| Zhang, Henan Medical Research, 2020 | Yes | Yes | Partial^b^ | Yes | Partial^d^ | Partial^i^ | Partial^j^ | Yes | N/A | N/A | N/A | N/A |
| Li, Yang Sheng Bao Jian, 2020 | Yes | Yes | Partial^b^ | No | Partial^d^ | Yes | Yes | Partial^k^ | N/A | N/A | N/A | N/A |
| Li, Today Nurse 2020 | Yes | Yes | No | Yes | No | Yes | Partial^j^ | Partial^l^ | Partial^n^ | N/A | N/A | N/A |
| Wei, Chinese Community Doctors, 2021 | Yes | Yes | Partial^c^ | Yes | Yes | Yes | Yes | Partial^m^ | N/A | N/A | N/A | N/A |
| Huang, Journal of Chengdu Medical College, 2020 | Yes | Yes | Partial^c^ | Yes | Yes | Yes | Yes | Yes | N/A | N/A | Yes | No |

^a^The authors do not describe where participants were asked to complete the writing intervention; ^b^The authors state the approximate intervention duration but they don't describe whether participants were asked to write specific amount; ^c^Materials are described but it is unclear how to access them; ^d^The authors describe some personnel but their intervention-specific training is unclear; ^e^The name or description of the online intervention platform is not provided; ^f^The authors state that participants were sent a link but the platform and physical space where participants were allowed to complete the intervention was not described; ^g^The physical space where participants were allowed to complete the intervention was not described; ^h^The order of the intervention components was not specified; ^i^It is unclear whether the intervention was delivered individually or in a group; ^j^The authors state that the intervention was delivered in a hospital but it's unclear where in the hospital it was delivered; ^k^The frequency of the intervention is not described; ^l^The frequency and duration of the relaxation and group therapy aspects of the intervention were not described; ^m^The duration of each intervention session and entire intervention period were not described; ^n^The authors state that younger children were educated about COVID-19 with images and stories. No further description is provided

**Appendix 4. Trials Excluded Because Participants Infected or Isolated, Follow-up Less than a Week, or non-Randomised**

**Additional Information also available at https://www.depressd.ca/research-question-3-intervention**

***Trial Characteristics (N = 73)***

| Study Details | Participants | | Country | | Intervention | | Comparator | Dates of Data Collection | N Intervention (Analyzed) | N Comparator (Analyzed) | Outcome Domain(s) | Mean (SD) Age | % Female or Women |
| --- | --- | --- | --- | --- | --- | --- | --- | --- | --- | --- | --- | --- | --- |
| Trials conducted among people with acute stress; people hospitalized or isolated due to COVID-19 infection or exposure (N=59; Verified=1, Unverified=58) | | | | | | | | | | | | | |
| Liu, Complementary Therapies in Clinical Practice, 2020  DOI: 10.1016/j.ctcp.2020.101132 | Patients hospitalized with confirmed COVID-19 | | China | | Progressive muscle relaxation and deep breathing 20-30 minutes per day for 5 consecutive days | | Standard care | 01/2020-02/2020 | 25 | 26 | Anxiety | 50 (13) | 45 |
| Liu, Medical Journal of Air Force, 2020  DOI:10.3969/j.issn.2095-3402.2020.03.024 | Patients hospitalized between 03/02/2020 to 30/03/2020 due to COVID-19 | | China | | Individualized psychotherapy plus group music therapy and physical exercise | | Standard care including general education of the virus and dietary regulation | NR | 75 | 75 | Anxiety, Depression | Intervention 54 (7) Comparator 50 (7) | 59 |
| Wu, Journal of Nursing Science, 2020  DOI: 10.3870/j.issn.1001-4152.2020.13.074 | Female aged 19-70 with oxyhemoglobin saturation greater than 0.93; with no other serious cardiopulmonary diseases other than COVID-19 infection | | China | | TCM emotional therapy focusing on positive emotions, meditation, breathing | | Standard care | NR | 100 | 100 | Anxiety, Depression | 50 (11) | 100 |
| Deng, Journal of Yangtze University, 2020  DOI:10.16772/j.cnki.1673-1409.2020.04.017 | Mild to moderate COVID-19 infected patients with no other underlying diseases | | China | | Standard psychological nursing plus personalized psychological nursing and peer-support group, including online psychological counselling 4x over 2 weeks | | Standard psychological nursing, including education about COVID-19 | NR | 30 | 30 | Anxiety, Depression | NR | 45 |
| Gharaati Sotoudeh, Iranian Journal of Psychiatry, 2020  DOI:   10.18502/ijps.v15i3.3812 | Adult COVID-19 patients admitted to Ziaeian general hospital | | Iran | | 4x 60 minute therapy sessions within 1 month, administered by 2 clinical psychologists, focusing on empathy, adjustment, responsibility, and spirituality | | Standard treatment and psychotherapy | 05/2020-06/2020 | 14 | 16 | Mental Health Function, Depression, Anxiety, Stress | Range:  18-65 | 53 |
| Hu, Jia You Yun Bao, 2020  Chinese Library Classification Number R47 | People hospitalized for more than 2 weeks from 02/2020 to 03/2020; with literacy and clear consciousness; without mental diseases or other systematic disorders | | China | | Participants instructed to complete daily diary entries | | Standard care | NR | 19 | 19 | Anxiety, Depression | Intervention 42 (7) Comparator 41 (7) | 45 |
| Hui, Journal of International Psychiatry, 2020  DOI:10.13479/j.cnki.jip.2020.04.001 | Patients hospitalized from 01/2020 to 02/2020 | | China | | 2x per day, 14-day psychological crisis intervention of the Neuman health system model | | 2x per day, 14-day standard psychological care | NR | 36 | 36 | Anxiety, Depression | Intervention 57 (8) Comparator 58 (8) | 46 |
| Zha, Journal of Hubei Minzu University- Medical Edition, 2020  Chinese Library Article Number: 2096-7578(2020)03-0102-03 | Literate adults hospitalized from 02/2020 to 03/2020 who know how to use WeChat correctly and have no prior mental disorder, neurological disorder, or other severe medical conditions | | China | | "Humanized Nursing" with the involvement of patients' family members through WeChat groups for promoting psychological care and arranging daily video chat for patients with family members | | Standard care | NR | 20 | 20 | Anxiety, Depression | Intervention 38 (2) Comparator 39 (2) | 40 |
| Chen, Nursing of Integrated Traditional Chinese and Western Medicine, 2020  DOI: 10.11997/nitcwm.202008027 | People hospitalized due to COVID-19 from 02/2020 to 03/2020, only adults over 18 | | China | | Baduanjin exercise twice a day | | Standard care | NR | 45 | 14 | Anxiety | Intervention 61 (16) Comparator 60 (15) | 59 |
| Cao,  Chinese Community Doctors, 2020  DOI:  10.3969/j.issn.1007-614x.2020.27.067 | Patients aged 41 to 79 who have no vital organ failure and loss of consciousness; and have no malignant tumor growth, were hospitalized from 01/2020 to 03/2020 | | China | | Humanistic care nursing: trained nursing team coordinates nutrition plan, individualized pulmonary recovery plan, and attends to patients' emotions | | Standard care | NR | 68 | 68 | Anxiety, Depression | Intervention 59 (11) Comparator 60 (12) | 29 |
| Li,  Chinese Community Doctors, 2020  DOI:  10.3969/j.issn.1007-614x.2020.27.070 | Patients hospitalized from 01/2020 to 04/2020 | | China | | "High quality nursing" including psychological and dietary intervention | | Standard care | NR | 58 | 57 | Mental Health Function, Depression, Anxiety | Intervention 54 (3) Comparator 53 (3) | 47 |
| Chen, Chinese Journal of Convalescent Medicine, 2020  DOI:10.13517/j.cnki.ccm.2020.11.006 | People hospitalized from 02/2020 to 03/2020 due to COVID-19 | | China | | 3-week Baduanjin exercise, 10 times per week, 2 times per day | | Standard care | NR | 14 | 15 | Anxiety, Depression | Intervention 68 (11) Comparator 69 (11) | 55 |
| Nie, Home Medicine, 2020  Chinese Library Article Number: 1671-4954(2020)04-0340-01 | Severely infected patients hospitalized from 02/2020 and 03/2020 | | China | | Humanistic intensive care including attention to negative emotions, providing proper diets, education on COVID-19 and protecting patients' privacy | | Standard intensive care | NR | 15 | 15 | Anxiety | Intervention 36 (8) Comparator 36 (3) | 43 |
| Zhang, Chinese Journal of Nursing, 2020  DOI:10.3761/j.issn.0254-1769.2020.S1.229 | Patients who were admitted to hospital from 22/01/2020 to 10/02/2020, with no serious complication | | China | | 10 days, ADOPT mode intervention: personalized guidance and issued questionnaires; organizing patients to watch educational videos and discuss effective plans every other day; evaluate the effect together | | Standard nursing, including health education, diet and medication guidance. | NR | 12 | 12 | Anxiety | 46 (3) | 46 |
| Pan, Yi Yao Qian Yan, 2020  Chinese Library Article Number: 2095-1752(2020)16-0184-02 | Patients hospitalized from 01/2020 to 03/2020 | | China | | Routine care plus psychological interventions. Different intervention methods were adopted for patients with different symptoms. 1.For panic patients: select more experienced nursing staff to take care of them during the rescue process. 2.For patients with dysphoria: keep absolutely patient and sympathetic; play appropriate music. 3.For patients with depression: analyze the origin of their pessimistic mood and have a targeted communication | | Routine Care | NR | 30 | 30 | Anxiety, Depression, Mental Health Function | NR | 45 |
| Dong, Yi Yao Qian Yan, 2020  Chinese Library Article Number: 2095-1752(2020)18-0152-02 | Patients with complete medical record, hospitalized from 02/2020 to 03/2020 | | China | | Routine nursing plus humanistic care, including sanitary improvement of the ward; educating patients with COVID-19 related knowledge; and using verbal encouragement to decrease patients' level of anxiety and loneliness; encouraged to communicate with their families via telephone and video | | Routine nursing, including nutrition guidance and symptom monitoring | NR | 46 | 46 | Anxiety, Mental Health Function | Intervention 42 (7) Comparator 42 (8) | 45 |
| Zhu, Infectious Disease Information, 2020  DOI: 10.3969/j.issn.1007-8134.2020.05.017 | Patients who were hospitalized from 02/2020 to 03/2020 | | China | | 9 days, One on one health education; medication guidance and vital signs monitoring; instruct patients to do breathing exercises twice a day to relax mind | | Standard nursing, including explaining COVID-19 related knowledge | NR | 40 | 40 | Anxiety, Depression | Intervention 36 (11) Comparator 36 (13) | 43 |
| Zhan, Yin Shi Bao Jian, 2020  Chinese Library Article Number: 2095-8439(2020)7-0134-01 | COVID-19 infected patients who were hospitalized from 25/01/2019 to 31/01/2020 and 01/02/2020 to 07/02/2020 | | China | | Standard care plus psychological intervention, including enhanced communication with patients; educating patients with COVID-19 related information; assessing patients' psychological status; and attending to patients' emotions | | Standard care | NR | 32 | 32 | Anxiety, Depression | Intervention 44 (3) Comparator 43 (2) | 52 |
| Jiang, Jia You Yun Bao, 2020  Chinese Library Classification Number: R395.1 | Mild COVID-19 patients with anxiety and depression, hospitalized from 25/02/2020 to 15/03/2020 | | China | | Standard nursing plus psychological care, including health education, communication with patients, playing positive movies and soft songs, mindfulness therapy | | Standard nursing | NR | 43 | 43 | Anxiety, Depression | NR | NR |
| Guo, Yin Shi Bao Jian, 2020  Chinese Library Article Number: 2095-8439(2020)7 -0223- 02 | Patients hospitalized at the particular hospital from 02/2020 to 03/2020 | | China | | Psychological nursing, including evaluating patients' psychological status, preventing patients from having negative emotions, and encouraging family support | | Standard nursing | NR | 30 | 30 | Depression, Anxiety | Intervention median 54 (3) Comparator median 53 (3) | 45 |
| Li,  Frontiers in Psychiatry, 2020  DOI: 10.3389/fpsyt.2020.580827 | Patients with mild symptoms without previously diagnosed depression or currently taking medication and had no prior cognitive dysfunction or experienced another major stressful event | | China | | Routine treatment plus 30-min CBT in the morning on a daily basis, including cognitive intervention that helps correcting patients' misconceptions in regard to COVID-19 information and management, relaxation techniques training, problem-solving training, and social support encouragement | | Routine treatment, including antiviral treatment, symptomatic treatment of fever, and nursing care | 02/2020 - 03/2020 | 47 | 46 | Depression, Anxiety, Stress | Intervention 48 (12) Comparator 47 (11) | 65 |
| Chen, Home Medcine, 2020  Chinese Library Article Number: 1671-4954(2020)04-0319-01 | Patients admitted to the Infectious Diseases Department during COVID-19 | | China | | Standard nursing plus humanized nursing (psychological care; health education; relaxation training therapy) | | Standard nursing | NR | 40 | 40 | Anxiety | NR | NR |
| Zhang, Practical Clinical Medicine, 2020  DOI:10.13764/j.cnkil.csy.2020.08.027 | Patients hospitalized from 15/02/2020 to 15/03/2020 | | China | | Fitness Qigong (Yijinjing): group training: one week, 3 days a week, twice a day, 60 minutes once; personal practice according to their own physical conditions; Chinese medicine emotion nursing: from 4pm to 5pm everyday; ear acupoint bean-pressing: 10-15 times a day, 3-5 minutes once | | Routine nursing, including symptom monitoring, medicine and nutrition guidance | NR | 14 | 14 | Anxiety, Depression | 50 (4) | 39 |
| Fan, Ke Xue Yang Sheng, 2020  Chinese Library Classification Number: R47 | Patient hospitalized from 01/2020 to 03/2020 | | China | | Systematic nursing, including psychological nursing to alleviate patients' negative emotions; rehabilitation training to enhance immune system; increased ward rounds to monitor changes in patients' symptoms; and nutrition guidance | | Routine nursing, including health education, food guidance, rehabilitation training | NR | 50 | 50 | Depression, Anxiety | Intervention 44 (6) Comparator 46 (6) | 48 |
| Gao, Ke Xue Yang Sheng, 2020  Chinese Library Classification Number: R47 | Patients registered at the hospital from 01/2020 to 02/2020 | | China | | Psychological intervention, including attending to patients' emotions and providing targeted psychological leading; weekly psychological counselling by professional counsellor; and providing improved nursing service | | Standard care, including monitoring patients' vital signs and medication guidance | NR | 15 | 15 | Depression, Anxiety | Intervention 54 (4) Comparator 54 (4) | 43 |
| Wang, Chinese Community Doctors, 2020  DOI:  10.3969/j.issn.1007-614x.2020.33.082 | People had close contact with COVID-19 infected patients and people suspected to have COVID-19 infection with ZSAS > 40, or ZSDS > 41, or PSQI > 16 | | China | | 30min daily personalized psychological intervention tailored for three types of psychological response, including providing COVID-19 related health education for people with anxiety symptoms; changing the negative cognition of people with depression symptoms; and educating and providing soft music for people with nervosism symptoms | | Conventional psychological intervention, including health education and psychological counselling | NR | 40 | 39 | Depression, Anxiety | Intervention 56 (14) Comparator 58 (13) | 56 |
| Shi, World Latest Medicine Information, 2020  DOI:10.3969/j.issn.1671-3141.2020.87.123 | People hospitalized from 01/2020 to 03/2020 | | China | | Daily psychoeducation focused on emotional regulation; psychological assessment of patients on the 1st, 7th and 14th day after admission; daily TCM emotional therapy; according to the constitution type to give the corresponding diet therapy | | Standard nursing care | 01/2020 - 03/2020 | 30 | 30 | Depression, Anxiety | Intervention 32 (2) Comparator 35 (3) | 48 |
| Ma, Nursing Research, 2020  Chinese Library Article Number: 1006― 6845(2020)8― 0136― 01 | Patients hospitalized from 12/02/2020 to 18/03/2020 | | China | | Standard nursing plus improved ward environment and individualized humanistic intervention focused on alleviating patients' negative affects | | Standard care | NR | 130 | 130 | Depression | 64 (NR) | 42 |
| Zhu, Practical Clinical Medicine, 2020  Chinese Library Article Number: 1009-8194(2020)10-0076-03 | Patients with no psychiatric disorder or low rates of survival | | China | | Standard nursing plus individualized daily Acceptance and Commitment Therapy (ACT), with each session lasting from 15-30min for a total of 5-6 sessions, content including helping patients accepting the influences brought by COVID-19, isolating from negative cognitions, and focusing on the present positivities and the self | | Standard nursing | NR | 46 | 46 | Anxiety, Depression | Intervention 65 (NR) Comparator 67 (NR) | 45 |
| Liu,  Yin Shi Bao Jian, 2020  Chinese Library Article Number: 2095-8439(2020)9-0126-01 | Mildly infected patients with blood oxygen saturation greater than 93% and breathing rate less than 24/min; with no psychiatric disorder or other comorbidity; not currently in gestation period | | China | | Standard nursing plus psychological nursing and humanistic nursing, focusing on monitoring and stabilizing patients' affects and helping them accepting their negative emotions. | | Standard nursing | NR | 22 | 22 | Anxiety, Depression | 49 (3) | 52 |
| Liu, Yin Shi Bao Jian, 2020  Chinese. Library Article Number: 2095-8439(2020)-0170-02 | Patients hospitalized from 20/01/2020 to 26/02/2020 | | China | | Standard nursing plus humanistic care, including nutrition guidance, providing comfortable ward environment, and psychological nursing to alleviate patients' negative emotions and cognitions. | | Standard nursing | NR | 20 | 20 | Anxiety, Depression | Intervention 54 (8) Comparator 45 (7) | 60 |
| Pan, Yi Yao Qian Yan, 2020  Chinese. Library Article Number: 2095-8439(2020)9-0241-01 | Patients hospitalized from 01/2020 to 04/2020 | | China | | Psychological nursing, including building comfortable environment, stabilizing patients' mood, educating patients about COVID-19 knowledge, and alleviating patients' negative affects | | Standard nursing | NR | 16 | 16 | Anxiety, Depression | Intervention 52 (5) Comparator 53 (6) | 44 |
| Wang, Jia You Yun Bao, 2020  Chinese Library Classification Number: R47 | Patients with no prior mental illness history | | China | | Traditional Chinese music intervention | | Standard care | NR | 32 | 33 | Mental Health Function, Depression, Anxiety | Intervention 47(11) Comparator 47 (12) | 57 |
| Shi, Smart Healthcare, 2020  DOI:10.19335/j.cnki.2096-1219.2020.20.054 | Adult ICU patients without COVID-19 infection; and have no pre-existing mental disorders, insomnia, or severe comorbidity | | China | | Psychological care, including educating patients with COVID-19 related information and helping patients relax both physically and psychologically | | Standard care | 01/2020 - 04/2020 | 30 | 30 | Anxiety | Intervention 52 (10) Comparator 51 (9) | 45 |
| Liu, Yin Shi Bao Jian, 2020  Chinese. Library Article Number: 2095-8439(2020)-001002 | COVID-19 isolates in 02/2020 with no malignant tumor or mental diseases | | China | | Improved isolation environment with psychological guidance | | Standard care | NR | 20 | 20 | Anxiety, Depression | Intervention 49 (4) Comparator 48 (4) | 43 |
| Lin, Health Vocational Education, 2020  Chinese. Library Article Number: 1671-1246(2020)20-0149-02 | Patients with body temperature greater than 37.3 degree celsius and an epidemiological history; have coughing and diarrhea symptoms admitted to hospital from 23/01/2020 to 16/02/2020, | | China | | COVID-19 and fever-related education including brochures and one-on-one health counseling | | Standard care | NR | 50 | 50 | Anxiety, Depression | Intervention 39 (10) Comparator 39 (8) | 48 |
| Li,  Home Medicine, 2020  Chinese Library Article Number: 1671-4954(2020)05-0259-02 | Patients hospitalized from 24/01/2020 to 24/02/2020 | | China | | Psychological intervention, including attending to patients' needs and emotions; providing music intervention; form patient support WeChat group and WeChat counselling, where nursing staff chat with patients twice daily with each round greater than 30min; and arrange professional psychology counsellor to have one-on-one counselling with patients | | Routine nursing, including diet and medication guidance | NR | 100 | 100 | Anxiety, Depression | Intervention 41(13) Comparator 43(13) | 35 |
| Wei, Journal of Zhejiang University-SCIENCE B (Biomedicine & Biotechnology), 2020  https://doi.org/10.1631/jzus.B2010013 | Laboratory confirmed COVID-19 adult patients in the isolation ward with PHQ-9 and GAD-7 ≥5 and ≤ 15, completed at least junior middle school, no suicidal ideation or antipsychotic use | | China | | Self-help intervention containing four main components: breath relaxation training, mindfulness (body scan), “refuge” skills, and butterfly hug method. Participants instructed to follow 50 min recordings daily for 2 weeks. | | Daily supportive care | 02/2020-02/2020 | 13 | 13 | Anxiety, Depression | Intervention 41(14)  Comparator 49(10) | 38 |
| Wu, Ke Xue Yang Sheng, 2020  Chinese Library Classification Number: R47 | Patients registered at the hospital between 2019/12 and 2020/02 | | China | | Psychological intervention, including informing patients with COVID-19 related knowledge; providing focused psychological leading; creating comfortable environment for patients; and regular monitoring of patients' vital signs | | Standard care, including regular disinfection, routine monitoring of patients' temperature, and educating patients with COVID-19 related self-care information | NR | 250 | 250 | Anxiety, Depression, Mental Health Function | Intervention 51(1)  Comparator  48(2) | 49 |
| Xu, Yin Shi Bao Jian, 2020  Chinese Library Article Number: 2095-8439(2020)8-0161-01 | Staff isolated between 2020/02 and 2020/04 | | China | | Proper meal provided by nutritionists which includes post-meal fruits | | Standard meal | NR | 79 | 79 | Anxiety, Depression | 26 (7) | 24 |
| Huang,  Nursing research, 2020  Chinese Library Article Number: 1006-6845(2020)8-0092-02 | Patients hospitalized at the particular hospital | | China | | Health education and psychological nursing during and after treatment focused on eliminating patients' negative affect | | Standard nursing | NR | 37 | 36 | Anxiety, Depression | 56 (NR) | 53 |
| Wang, Laboratory Medicine and Clinic, 2020  DOI: 10.3969/j.issn.1672-9455.2020.23.033 | Severely infected adult patients with no recent major life events taken place, with no malignant tumor or prior psychiatric disorder, or taking anti-depressants | | China | | Routine nursing plus daily empathic psychological intervention for 20-30min, including empathic listening, transposition thinking, information organizing, and feedback exchanging. | | Standard nursing, including health education, disinfection, and food guidance | NR | 20 | 20 | Anxiety, Depression | Intervention 46 (7)  Comparator  46 (7) | 45 |
| Liu, Modern Practical Medicine, 2020  DOI: 10.3969/j.issn.1671-0800.2020.11.044 | Adult patients aged 18-60 with no cognitive dysfunction, mental or psychiatric disorder, or severe underlying diseases | | China | | 90-120min daily art therapy for 10 days, focusing on building confidence, strengthening self-conception, enhancing self-efficacy, and looking forward to the future | | Standard nursing, including health education and rehabilitation exercise | NR | 25 | 25 | Anxiety, Depression, Mental Health Function | Intervention 43(8)  Comparator  44(10) | 42 |
| Wei, Gansu Science and Technology, 2020  Chinese Library Classification Number: R473.5 | Medical staff in their medical observation period after contacting with patients with coronavirus infection | | China | | Daily psychological counselling via WeChat group for 14 days | | Routine activity | 02/2020-02/2020 | 35 | 35 | Anxiety, Depression | Intervention 32 (2)  Comparator  31(2) | 73 |
| Xu, Yin Shi Bao Jian, 2020  Chinese Library Article Number: 2095-8439(2020)-0210-02 | Patients hospitalized between 2020/2/5 and 2020/4/5 | | China | | Routine nursing plus humanistic care focusing on improving patients' negative emotions, including providing a comfortable ward environment, psychological care targeting patients' needs, and spiritual support by reminding patients of their family | | Routine nursing | NR | 40 | 40 | Anxiety, Depression | Intervention 60 (7)  Comparator  59 (7) | 43 |
| Wang, Yin Shi Bao Jian, 2020  Chinese Library Article Number: 2095-8439(2020)9-0142-02 | Patients hospitalized between 2020/02 and 2020/03 | | China | | Routine nursing plus targeted psychological nursing intervention, including providing the patients and their family about COVID-19 information, and teaching patients relaxation techniques and psychological regulation techniques | | Routine nursing, including enhanced monitoring of vital signs and support treatment, and providing stabilization of patients' internal environment | NR | 39 | 39 | Anxiety, Depression | Intervention 57 (4)  Comparator  56 (4) | 47 |
| Cai, World Latest Medicine Information, 2020  DOI: 10.3969/j.issn.1671-3141.2020.98.038 | Mildly infected or ordinary type patients aged 18-65, with no severe underlying diseases | | China | | General medicine intervention, including health education in regards to COVID-19; psychological intervention targeting at alleviating patients' negative emotions; medical intervention; nutrition guidance; and exercise instructions | | Standard nursing | NR | 30 | 30 | Anxiety, Depression | Intervention 43 (12)  Comparator  43 (11) | 45 |
| Si, Journal of Modern Medicine & Health, 2021  DOI: 10.3969/j.issn.1009-5519.2021.02.011 | Accompany for critically ill patients in the department of cardiology during COVID-19 | | China | | Targeted intervention, including delivering health education about COVID-19 and organizing family visit; rewards are provided for participants' family with high compliance to conducting COVID preventative measures | | Routine activity | NR | 65 | 45 | Anxiety, Depression | 45 (10) | 56 |
| Du, Laboratory Medicine and Clinic, 2021  DOI: 10.3969/j.issn.1672-9455.2021.02.036 | Mild covid-19 patients hospitalized between 2020/01 and 2020/03, no history of major disease, with clear consciousness | | China | | Routine psychological care and narrative nursing intervention. Narrative nursing was performed 3 times a week for a month, each time 20-30mins: guiding the patient to narrate; implementing eye closure desensitization method, music and dance relaxation method to relieve insomnia; through other successful events experienced by mild COVID-19 patients to help patient to establish the confidence to overcome the disease. If the patient is willing, his/her family members can watch the whole process through WeChat video. | | Routine psychological care: improve patients' cognition of COVID-19; communicate and keep gentle nursing actions when conducting COVID-19 treatment. | NR | 33 | 34 | Anxiety | Intervention median: 35  Comparator  median: 36 | 49 |
| Cai, World Latest Medicine Information, 2020  DOI: 10.3969/j.issn.1671-3141.2020.97.147 | Mildly infected COVID patients and patients suspected to have COVID-19 between age 18 and 60 with complete medical records | | China | | Seamless management intervention, including forming nursing teams with trained staff; providing psychological nursing to the patients via WeChat platform; encouraging peer support via leisure activities; and organizing educative sessions about COVID-19 information | | Routine nursing, including vital sign monitoring and health education | NR | 80 | 80 | Anxiety, Depression | Intervention 53 (7)  Comparator  49 (7) | 48 |
| Zhang, Contemporary Medical Symposium, 2021  DOI: 2095-7629-(2021)01-0089-03 | Patients with mild and ordinary type of COVID-19, with no cognitive disorders. | | China | | Routine treatment plus the combination of Fei Yan Yi Hao Fang (TCM prescription for pneumonia) and shared decision-making psychological counselling to enhance patients' confidence in fighting against the disease. | | Routine treatment, including maintaining patients' internal homeostasis, providing nutrition support, routine screening, and medication support, and the prescription of Fei Yan Yi Hao Fang (traditional Chinese medicine targeting pneumonia). | NR | 106 | 100 | Anxiety | Intervention 35 (5)  Comparator  36 (5) | 42 |
| Wang, Modern Nurse, 2021  DOI: 10.19793/j.cnki.1006-6411.2021.06.060 | Patients from the fever clinic who needed to remain in hospital under observation and have two nucleic acid testings, with no major diseases | | China | | Mental intervention, including Mindfulness-Based Stress Reduction, focused on helping patients relax and meeting their psychological needs, the intervention was delivered every two hours until the patient left the hospital, each session lasted 10-20min. | | Routine care and health education | NR | 55 | 55 | Anxiety | Intervention 34 (13)  Comparator  32 (12) | 51 |
| Min, Health Guide, 2020  Chinese Library Article Number: 1006-6845(2020)12-0009-02 | Quarantined people who were waiting for their diagnosis of coronavirus infection | | China | | Routine psychological care plus Psybot psychological intervention, which provided COVID-related information, stress-reduction games, and psychological counselling on demand. | | Daily routine psychological care targeted at decreasing participants' anxiety and fear. | NR | 45 | 45 | Anxiety, Depression | Intervention 39 (9)  Comparator  39 (8) | 36 |
| Shaygan, BMC Psychiatry, 2021  DOI: 10.1186/s12888-021-03085-6 | Adult patients diagnosed with mild-to-moderate or severe COVID-19 and had been hospitalized during the past 48 h; with no previous experience of quarantine, no history of psychiatric disorders or taking psychiatric medications. | | Iran | | Online multimedia psycho-educational intervention, of which consisted of 14 daily modules that were based on cognitive-behavioural techniques, stress management techniques, mindfulness-based stress reduction and positive psychotherapy; participants were required to complete 1 module per day which was designed to be 60 min in total; WhatsApp was used to deliver daily multimedia psycho-educational contents to the patients between 9 AM and 9 PM with approximately two-hour interval. | | Participants were offered the opportunity to receive telephone-based counselling from the psychological team if needed. | NR | 26 | 22 | Mental Health Function, Stress | 37 (12) | 44 |
| Ozlu, Perspectives in Psychiatric Care, 2020  DOI: 10.1111/ppc.12750 | Adult patients who were receiving treatment in the clinic with no visual or hearing impairments | | Turkey | | 20-30min progressive muscle relaxation exercises which were delivered on TV via CDs; participants performed the exercises twice a day for 5 days. | | Routine care | 05/2020-08/2020 | 33 | 34 | Anxiety | Intervention 36 (12)  Comparator  33 (12) | 45 |
| Wang, Medical Diet and Health, 2021  Chinese Library Article Number: 2096-5249(2021)03-0123-03 | Patients of suspected new types of coronary pneumonia, with HAMA ≥ 14 and number of hospitalized days >14d | | China | | Psychological intervention, including having 15-30min interviews with patients with anxiety; telephone follow-ups every 2 weeks for patients who left the hospital; daily music therapy for 30min to help patients maintaining positive moods; cognitive therapy to help patients forming the correct perception of COVID-19; and mindfulness-based stress reduction to help patients building confidence and alleviating negative emotions. | | Routine medication and psychological care | NR | 25 | 25 | Anxiety | Intervention 40 (8)  Comparator  44 (7) | 36 |
| Yang, World Latest Medicine Information, 2021  DOI: 10.3969/j.issn.1671-3141.2021.4.142 | Patients with mild coronavirus infection, with no severe cardiac, hepatic, or renal functional diseases, and no severe neurological diseases. | | China | | Traditional Chinese medicine (TCM) syndrome differentiation nursing methods which lasted for 1 month, combined with acupoint massage, the duration for each acupoint lasted 2min, and the message was performed 2-3 daily. | | Routine nursing | NR | 44 | 44 | Anxiety, Depression | 49 (0) | 33 |
| Non-randomised controlled trials (N=4) | | | | | | | | | | | | | |
| Weis, Couns Pychother Res. 2020  http://doi.org/10.1002/capr.12375 | Undergraduates enrolled in one of two psychology research classes at a midwestern university | | USA | | Four weekly Koru Mindfulness sessions (75 min) through group video conference and encouraged daily individual 10 minute practice sessions | | Waitlist | NR | 16 | 16 | Stress, Anxiety, Mental health function | 20 (1) | 78 |
| Kubo, PsyArXiv Preprint  http://doi.org/10.31234/osf.io/eb6yz | Students from the first grade to third grade at one junior high school | | Japan | | 50min class intervention consisting of a 15min stress reflection session and 35min psychoeducation session | | 1) An announcement about COVID-19 was made for the entire second grade at a term-end rally  2) Waitlist | 07/2020-08/2020 | 72 | 1) 92  2) 84 | Depression, Anxiety, Mental health function | NR | 48 |
| Liu, JMIR Mental Health, 2020  http://doi.org/10.2196/23917 | Mothers with 3-7 year-old children with autism | | China | | Joint Attention, Symbolic Play, Engagement, and Regulation (JASPER) online course (45-60 min 2x per week for 12 weeks), weekly online Q&A sessions, and online psychological counseling course (45-60 min, 6x over 12 weeks) | | Mothers were provided with an electronic manual "108 Strategies to Overcome the Pandemic at Home" and a home training plan for children (homework check-ins weekly for 12 weeks) | 01/2020-06/2020 | 65 | 60 | Anxiety, Depression, Stress | 33 (4) | 100 |
| Liu, Journal of Beijing Sport University, 2020  https://www.doi.org/10.19582/j.cnki.11-3785/g8.2020.03.009 | NR | | China | | 5-week, 3 times per week, online group Tai Chi intervention | | 1) 5-week, 3 times per week online physical fitness intervention  2) No intervention | NR | 266 | 1) 265  2) 258 | Depression, Anxiety | NR | NR |
| Brief trials without outcome assessment a week or longer after intervention initiation or trials where it was not clear when the outcome was assessed, and no author contact information was available (N= 10) | | | | | | | | | | | | | |
| [Cantarero,   Social Psychological and Personality Science,  2020  https://doi.org/10.31234/osf.io/pyhce](https://doi.org/10.31234/osf.io/pyhce) | | Mturk workers | | Mixed; countries not specified | Participants were asked to write about when during COVID-19 they have felt a sense of autonomy, sense of relatedness, and sense of competence | Participants wrote about their favorite colour | | NR | NR | NR | Stress, Quality of life | 37 (12) | 36 |
| Pizzoli, JMIR Mental Health, 2020  https://doi.org/10.2196/preprints.22757 | | Adult Italian-speakers | | Italy | Participants heard one 7-minute recorded voice guiding the regulation of breathing frequency, with the aim of making every breath cycle the same length | 1) Participants listened to one 7-minute audio clip with a voice that guiding a body-scan meditation  2) Participants were presented with one prerecorded audio clip of natural sounds (rain, water sounds) | | 05/2020-NR | 77 | 1) 76  2) 87 | Stress | 40 (15) | 72 |
| Hu, PsyArXiv, Preprint  https://doi.org/10.31234/osf.io/mc57s | | COVID-19 essential workers, i.e., medical staff and police officers | | China | Participants received a daily online survey at 20:00pm for five days which prompted them to watch a 2 minute video clip of natural scenes | Participants received a daily online survey at 20:00pm for five days which prompted them to watch a 2 minute video clip of urban scenes | | NR | 35 | 31 | Mental health functions | 36 (9) | 35 |
| Ding, Psychiatria Danubina, 2020  https://doi.org/10.24869/psyd.2020.527 | | Non-graduating (age 12-18) Chinese middle school students with an anxiety score greater than 50 points | | China | Routine health education plus 8 weeks of model 328-based exercise program including encouraged weekly online live peer-education seminars and aerobics 2x per day, 3 days per week scheduled every other day, with each exercise lasting 10min (guided by a recording) | Routine health education, including diet and nutrition, lifestyle habits, physical exercise, mental health, and pandemic-related knowledge. | | 02/2020-04/2020 | 70 | 71 | Depression, Anxiety | 15 (2) | 44 |
| [Zhang,  Nursing Research,  2020   https://kns.cnki.net/kcms/detail/detail.asp xdbcode=CJFD&dbname=CJFDLAST2020 &filename=NFHL202006012&v=zP%25 mmd2FU%25mmd2Bcdb0mLYUD9U 2rsUdjs34q7tcOMblTUFhq3fwflLpbrl kvTX3wQzVfXenqxB](https://kns.cnki.net/kcms/detail/detail.aspx?dbcode=CJFD&dbname=CJFDLAST2020&filename=NFHL202006012&v=zP%25mmd2FU%25mmd2Bcdb0mLYUD9U2rsUdjs34q7tcOMblTUFhq3fwflLpbrlkvTX3wQzVfXenqxB) | | Nurses from the department specialized in treating coronavirus infection | | China | Nursing narrative intervention, including listening to the nurses' experiences to understand their affects and attitudes; providing positive guidance and acknowledgement of the nurses' value and contributions; organizing regular psychology lectures to increase nurses' wellbeing; incorporating sandplay therapy to help nurses expressing emotions; increasing nurses' positivity by rational emotive therapy | Standard care with psychological guidance by the head nurse | | NR | 37 | 35 | Depression, Anxiety | Intervention 26 (4)  Comparator 27 (3) | 100 |
| [Wang, Nursing Research, 2020  http://d.wanfangdata.com.cn/ periodical/ChlQZXJpb2RpY2 FsQ0hJTmV3UzIwMjEwMzA yEhF5c2Jqem4teDIwMjAzNT E0MxoINXZjY3NqOHY%3D](http://d.wanfangdata.com.cn/periodical/ChlQZXJpb2RpY2FsQ0hJTmV3UzIwMjEwMzAyEhF5c2Jqem4teDIwMjAzNTE0MxoINXZjY3NqOHY%3D) | | Perinatal mothers enrolled from 2020.2.1 to 2020.2.29 | | China | Individualized psychological intervention focused on alleviating patients' fear and anxiety | Standard nursing | | NR | 42 | 42 | Depression, Anxiety | 27 (5) | 100 |
| [Han,  Electronic Journal Of Practical Clinical Nursing Science,  2020   http://med.wanfangdata.com.cn/ Periodical/sylchlxdzzz](http://med.wanfangdata.com.cn/Periodical/sylchlxdzzz) | | Medical staff in their medical observation period after contacting with patients with coronavirus infection | | China | Daily psychological counselling via WeChat group for 14 days | Routine activity | | NR | 35 | 35 | Depression, Anxiety | Intervention 32 (2)  Comparator 31 (2) | 73 |
| [Peng, Yin Shi Bao Jian, 2020  http://d.wanfangdata.com.cn/ periodical/ChlQZXJpb2RpY2 FsQ0hJTmV3UzIwMjEwMzAyEg95aW 5zYmoyMDIwMzYxNDkaCDIyaHVhcm1j](http://d.wanfangdata.com.cn/periodical/ChlQZXJpb2RpY2FsQ0hJTmV3UzIwMjEwMzAyEg95aW5zYmoyMDIwMzYxNDkaCDIyaHVhcm1j) | | Nursing staff registered at the hospital of study | | China | Routine training plus resilience support and cognitive therapy, focusing on enhancing staff's self-affirmation and assisting staff forming the correct perception of their working environment, including providing semi-structured interviews to educate staff members about the preventiveness of COVID-19 in order to alleviate negative emotions | Routine training, including COVID-19 prevention and protection information session | | NR | 20 | 20 | Mental Health Function, Burnout | Intervention 38 (3)  Comparator 37 (4) | NR |
| Li, Qinghai Yi Yao Za Zhi, 2020  DOI NR | | Nurses working at the frontline against COVID-19 during 2020.1 and 2020.4 | | China | Mindfulness-based stress reduction, intervention focused on sensory and perceptual training, mindfulness-based breathing, body scan, and mindful meditation | Routine psychological care focused on alleviating participants' distress of getting infected and avoiding overworking | | NR | 48 | 48 | Depression, Anxiety | 29 (4) | 100 |
| Yu, Capital Food Medicine, 2021  DOI NR | | Lung cancer patients in stable condition at home after treatment, with Karnofsky score > 60, without mental disorder, cognitive dysfunction, hematological disease, renal and liver or cardiovascular diseases. | | China | Routine nursing plus health education, which focused on COVID as well as lung cancer-related information; the intervention was delivered via WeChat group | Routine nursing, including weekly telephone follow-ups | | NR | 47 | 47 | Depression, Anxiety, Mental health function | Intervention 47 (4)  Comparator 48 (4) | 41 |
